# Joint probability approach for prognostic prediction of conditional outcomes: application to quality of life in head and neck cancer survivors

**DOI:** 10.1101/2024.12.16.24319067

**Authors:** Mauricio Moreira-Soares, Erlend I. F. Fossen, Aritz Bilbao-Jayo, Aitor Almeida, Laura Lopez-Perez, Itziar Alonso, Maria Fernanda Cabrera-Umpierrez, Giuseppe Fico, Susanne Singer, Katherine J. Taylor, Andrew Ness, Steve Thomas, Miranda Pring, Lisa Licitra, Stefano Cavalieri, Arnoldo Frigessi, Marissa LeBlanc

**Author notes:** Corresponding author: Marissa LeBlanc, Norwegian Institute of Public Health, Lovisenberggata 8, Oslo, 0456, Norway.

## Abstract

**Background:** Conditional outcomes are outcomes defined only under specific circumstances. For example, *future* quality of life can only be ascertained when subjects are alive. In prognostic models involving conditional outcomes, a choice must be made on the precise target of prediction: one could target future quality of life, given that the individual is still alive (conditional) or target future quality of life jointly with the event of being alive (unconditional).We aim to (1) introduce a probabilistic framework for prognostic models for conditional outcomes, and (2) apply this framework to develop a prognostic model for quality of life *3 years* after diagnosis in head and neck cancer patients.

**Methods:** A joint probability framework was proposed for prognostic model development for a conditional outcome dependent on a post-baseline variable. Joint probability was estimated with conformal estimators. We included head and neck cancer patients alive with no evidence of disease *12 months* after diagnosis from the UK-based Head & Neck 5000 cohort (N=3572) and made predictions *3 years* after diagnosis. Predictors included clinical and demographic characteristics and longitudinal measurements of quality of life. External validation was performed in studies from Italy and Germany.

**Findings:** Of 3572 subjects, 400 (11.2%) were deceased by the time of prediction. Model performance was assessed for prediction of quality of life, both conditionally and jointly with survival. C-statistics ranged from 0.66 to 0.80 in internal and external validation, and the calibration curves showed reasonable calibration in external validation. An API and dashboard were developed.

**Interpretation:** Our probabilistic framework for conditional outcomes provides both joint and conditional predictions and thus the flexibility needed to answer different clinical questions. Our model had reasonable performance in external validation and has potential as a tool in long-term follow-up of quality of life in head and neck cancer patients.

**Funding:** The EU.

**Research in context:** *Evidence before this study:* We searched for “head and neck” AND “quality of life” AND (“prognostic prediction” OR “machine learning” OR “prediction model”) on PubMed for studies published up to September 2024 and found 45 results. The prognostic models developed in the identified publications either excluded subjects who died during follow up or imputed quality of life with 0 for subjects that died during follow up. None of these publications explicitly address the implications of conditioning on survival, which introduces a significant risk of bias and may lead to invalid interpretations. These issues are well known in biostatistics and epidemiology but are often overlooked among machine learning practitioners and data scientists working with health data. Furthermore, recent methodological studies, such as van der Goorbergh et al. 2022, have been raising awareness about the importance of predicting probabilities that are well calibrated and suitable for answering the predictive questions of interest. Taylor et al. 2019 have shown in a systematic review that health-related quality of life in head and neck cancer survivors can be severely impaired even 10 years after treatment. The scoping review by Alonso et al. 2021 highlights the need for the development of prediction models for supporting quality of life in cancer survivors: from the 67 studies included, 49% conduct parametric tests, 48% used regression models to identify prognostic factors, and only 3% (two studies) applied survival analysis and a non-linear method.

*Added value of this study:* This study makes an important *methodological contribution* that can generally be applied to prognostic modeling in patient populations that experience mortality but where survival is not the main target of prediction. to the best of our knowledge, this is the first time that this problem is tackled in the context of clinical prognostic models and successfully addressed with a sound statistical-based approach. In addition, our proposed solution is model agnostic and suitable for modern machine learning applications. The study makes an important *clinical contribution* for long-term follow up of head and neck cancer patients by developing a joint prognostic model for quality of life and survival. To the best of our knowledge, our model is the first joint model of long-term quality of life and survival in this patient population, with internal and external validation in European longitudinal studies of head and neck cancer patients.

*Implications of all the available evidence:* The probabilistic framework proposed can impact future development of clinical prediction models, by raising awareness and proposing a solution for a ubiquitous problem in the field. The joint model can be tailored to address different clinical needs, for example to identify patients who are both likely to survive and have low quality of life in the future, or to predict individual patient future quality of life, both conditional or unconditional on survival. The model should be validated further in different countries.

## Introduction

Conditional outcomes can only be measured under specific conditions. We focus on outcomes that happen in the future, together with a conditioning event that also happens in the future. For example, future health-related quality of life (HRQoL) is only measurable when a subject is still alive. In these situations, two variables are considered, both happening in the future. In prognostic models involving conditional outcomes, a choice must be made on the precise aim of the prediction: either one is interested in the conditional outcome, or in the unconditional (joint) one. For the HRQoL example, the former targets quality of life, given that the individual is alive (conditional); the latter targets quality of life jointly with the event of still being alive (unconditional). The predictive probability of the first case assumes that the person is alive, in the second case the life status of the individual is also evaluated as a random event that might occur or not. Emphasis is frequently placed on the conditional outcome(1), leading to predictions that condition on the occurrence of the post-baseline event. We argue that relying solely on conditional probability often fails to yield sufficiently informative predictions for decision-makers, who could be interested in the unconditional event. Because both occur in the future, it can be relevant to assess the probability that the outcome occurs, while incorporating the probability that the conditioning event also happens, instead of assuming this to be true. A common choice in prognostic prediction is to develop the model only in subjects for whom the conditional outcome is observed, often without acknowledging the conditionality(2,3). Some studies attempt to include all patients by imputing the conditional outcome for the censored cases(4), which introduces a significant risk of bias and may lead to invalid interpretations.

In clinical prognostic models, the aim is to answer relevant clinical questions, ideally in a probabilistic manner. Previous prognostic models for HRQoL in cancer patients have used survival to the time of prediction as an inclusion criterion, e.g.(5), and therefore estimate the conditional probability. This restricts the clinical question to “What is the probability of having a certain quality of life in the future, assuming the patient survives?” The alternative questions is “What is the probability of both surviving and having a certain quality of life in the future?” This question incorporates survival as a random event and considers that some patients are unlikely to survive in the first place. Estimating the joint probability of surviving and having a certain level of HRQoL therefore appears relevant.

While our study has a generic value, we concretely focus on head and neck cancer (HNC) patients who often endure a significant symptom burden because of the disease and/or the treatment including swallowing, speaking, and breathing difficulties(6), eating problems(7), pain(8), difficulties with social contacts(9), and loss of income(10). Typically, HNC patients experience a decline in HRQoL following the start of treatment compared to diagnosis, yet they tend to have improved HRQoL after treatment conclusion, usually within one year(11–13), though impairments can remain a problem for up to 10 years or longer(14). It is therefore of clinical interest to develop a prognostic model for HRQoL over the long term*, i.e*., in the years following end of treatment. Such predictions could help clinicians to identify patients at high risk for HRQoL decline and implement early interventions to mitigate this risk.

This study aims to (a) introduce a probabilistic framework for prognostic models for conditional outcomes that depend on post-baseline variables, with flexibility to address different clinical questions and (b) apply this framework to develop joint prognostic model for HRQoL and survival over the long term in HNC patients.

## Methods

### Probabilistic framework

For each individual *i* ∈ {1,… , *N*}, let *x_i_* = (*x*_l_,… , *x_p_*) be a random vector of *p* predictors measured at a certain baseline timepoint *t_O_*, and *y_i_* ∈ ℝ be a continuous outcome measured at a follow-up timepoint *t_f_*, where *t*_O_ < *t_f_* . Define the post-baseline event with the survival function given by

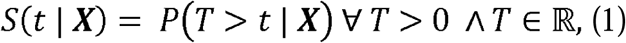

i.e., the probability that the time to event (*T*) is greater than some specific time t. The measurement of y at time *T=t* is conditionally dependent on survival to time *t*. We then define the joint probability of the continuous outcome being between the lower and upper thresholds (*h_L_, h_U_*) and survival to time *t*

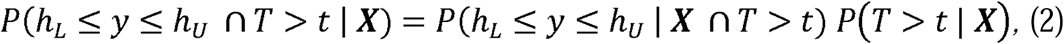

where the first term in the right-hand side of the equation is the conditional probability of measuring the continuous outcome between the thresholds given the survival time *T> t*, and the second term is the survival probability (note: this can be defined more generally by considering that the outcome *y* can belong to any given set *G* (*y* ∈ *G*) but for simplicity we defined *G*= *h_L_* ≤ *y* ≤ *h_U_* in Eq.(2) which is particular to the case example we explore in this paper.) To estimate the joint probability, we can develop two sub-models for (i) the conditional outcome

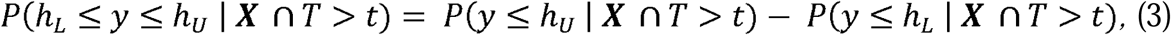

where we estimate the probability *P*(*y* ≤ h | ***X*** ⋂ *T* > *t*) for a generic threshold h so at inference time we can retrieve the probability in the desired interval by providing the specific lower and upper boundaries. And (ii) the model for estimating the probability for time to event

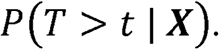

### Probability estimation

To obtain the probabilities in Eq (3), we use the model agnostic prediction interval estimator (MAPIE)(15) with cross-validation plus (CV+), which is a flexible estimator for predictive distributions of continuous outcomes. For (ii) any time-to-event method can be used, but the data in our case example present no censoring so we use LASSO logistic regression to estimate the survival probability to time t=2 years.

### Application of the framework to BD4QoL

#### Data sources

The Head and Neck 5000 study (HN5000) is a United Kingdom (UK)-based study of individuals with HNC. It is sponsored by the University Hospitals Bristol and Weston NHS Foundation Trust (UHBW) and conducted in collaboration with the University of Bristol(16,17). From 2011 to 2014, 5,511 participants were recruited at diagnosis from 76 centres. Over 200 variables were collected from diagnosis up to three years follow-up, encompassing clinical, demographic, HRQoL, physical, and mental health data. Survival data were obtained from participating sites and from NHS England. We accessed the HN5000 data for prediction model development via the BD4QoL European project (grant no. 875192). All studies collected HRQoL with the European Organisation for Research and Treatment of Cancer Core Questionnaire (EORTC QLQ-C30) and its head and neck cancer module (EORTC QLQ-H&N35)(18,19).

We screened for eligibility criteria (Supplementary Methods S1) to identify patients who were alive and with no evidence of disease at *12 months* after diagnosis (the first visit after end of treatment), resulting in N=3572 participants eligible for model development.

#### Outcome

The outcome is defined based on the Global Health Status /Quality of Life (GHS) measured with the respective scale in the European Organisation for Research and Treatment of Cancer Core Questionnaire (EORTC QLQ-C30)(18) at *3 years* after diagnosis. We will refer to GHS/QoL as simply QoL. The QoL is a pseudo-continuous scale that can take on eight possible values between 0 and 100, derived from two questions(20).

Using the proposed probabilistic framework we can make prognostic predictions for QoL. Clinically relevant questions may include “what is the probability that, 2 years from now, this patient is alive and with lower QoL than today?” or “if this patient survives the next two years, what is the probability that their QoL is above a certain value?”.

Below we show how to use the framework to model one particular target, i.e., the probability of being alive with a future QoL less than or equal to a given threshold. Here the time of prediction *t_f_* is at 3 years after diagnosis. In our data we have complete information (no censoring) for survival. Therefore, instead of modelling survival as a time to event, we model it as binary variable *S*(*t* = *t_f_*) = {0, 1} defined at the time of prediction *t_f_* that indicates if the patient was alive at prediction time or not. This makes the presentation easier, but more general situations are straightforward.

Applying our probabilistic framework, we define the outcome measurement as the QoL score *y* := *QoL*. Thus, Eq.(3) becomes

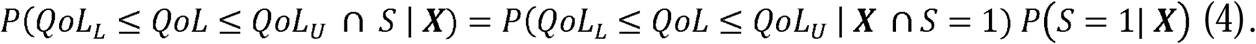

In this application, clinically significant deterioration in QoL is of interest, defined as a decrease of 10 points or more between *t_f_* and *t_O_*, hereafter referred as “QoL decline”. Therefore, we will obtain the QoL decline probability after the general model estimation for the probability in Eq.(4) by defining *QoL_L_* =0 and *QoL_U_* = *QoL_tO_*-10. Eq.(4) then becomes simply

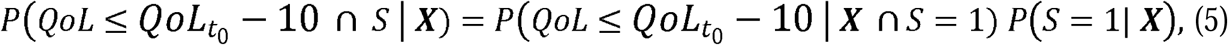

or, in terms of the complement, the probability of having a QoL decline or dying, *D*= *S^C^*

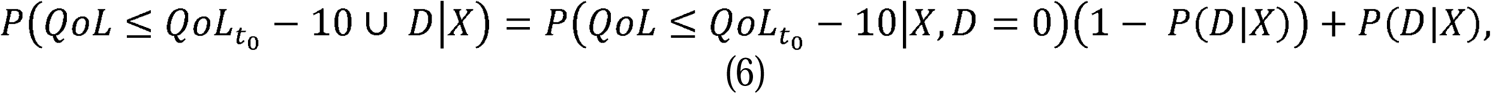

see Supplementary Methods S2 for complete derivation.

#### Candidate predictors

We developed models with two different sets of predictors:

1. An <u>extensive</u> set of 143 variables (171 parameters) to identify prognostic factors in an agnostic manner.
2. A <u>limited</u> subset of 43 variables (59 parameters)(21).

The extensive set included all available predictors in the data measured at diagnosis, at 4 months after diagnosis, and at the end of treatment at 12 months (i.e. measured at or before time 0 in our model). The limited set was chosen due to their known association with QoL(22) and their ready availability in the clinical setting. Variables include demographics information, clinical data, QoL questionnaires (e.g. EORTC QLQ-C30 and its module EORTC QLQ-H&N35(18)), and other psychometric information. Descriptions of the two lists of predictors can be found in the supplementary Table S1.

#### Sample size

The estimated sample sizes(23) required for developing the prognostic models are 1430 and 4249 subjects for 59 and 171 predictor parameters, respectively. The number of available subjects is 1631, which is adequate for the limited set but may increase the risk of overfitting for the extended set. To address this issue, we used an agnostic penalisation method. The detailed calculations can be found in the Supplementary Methods S3 and Tables S2 and S3.

### Statistical analysis

We used Apptainer with Python v3.10 for model development and Conda environment for exploratory analysis. Descriptive statistics were performed in R v4.2.3. The configuration files containing the full list of packages with versions and the code are available (https://github.com/ocbe-uio/BD4QoL). We used SHapley Additive exPlanations (SHAP) to estimate feature importance. We deployed models with an application programming interface (API) developed with FastAPI and a dashboard in HTML and JavaScript (https://github.com/ocbe-uio/BD4Predict). This report adheres to the TRIPOD checklist for transparent reporting(24) (supplementary Table S4).

### Missing data

Missing values in continuous and ordinal predictors were imputed with K-nearest neighbour (KNN) method (k=5 and uniform weights) and missing indicator method (MI) was used for categorical variables. We did not impute missing outcomes but a propensity scores model was estimated for evaluating model performance (Supplementary Methods S4). We only imputed variables that presented less than 50% missingness, the remaining were excluded from the analysis.

### Model development

We modelled the QoL score distribution with multivariable conformal models(25,26), to obtain the probability of QoL decline. The base estimators were LASSO linear regression and boosted trees. The predicted risk of death was modelled with LASSO logistic regression. Further details can be found in the Supplementary Methods S6.

### Internal validation

We performed bootstrap internal validation with 200 resamples, applying the .632+ estimator to obtain the C-statistics, R-squared, and the mean absolute error (MAE)(27). We used the .638 estimator(28) to obtain the observed-expected ratio (O/E), calibration-in-the-large (CIL), and calibration slope (CS). Calibration plots were estimated with local regression. See Supplementary Methods S7 for a complete description.

### External validation

External validation for the conformal model was performed with independent data from the University of Mainz (Germany) and the Istituto Nazionale dei Tumori (Italy). See Supplementary Methods S8 and S9 for more information about the data sources and Tables S11 and S12 for descriptive statistics.

## Results

### Participants

After screening the data for the eligibility criteria, 3572 subjects were included in the analyses (Figure 1). Survival status was available for all subjects, with 400 (11.2%) deceased at time of prediction (3 years after diagnosis). The QoL outcome was measured in 1662/3172 (52.4%) remaining alive.

**Figure 1.**
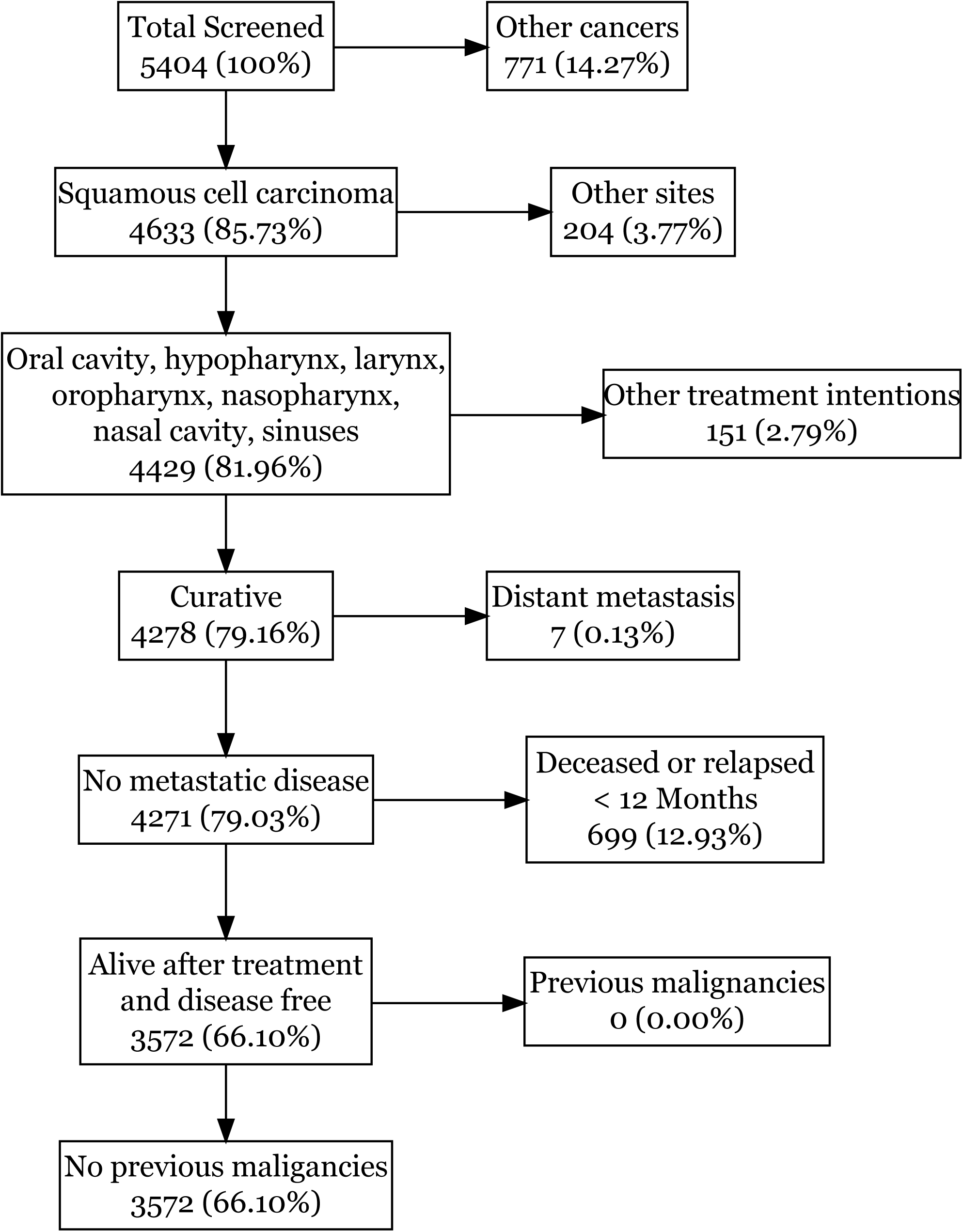
Flow of participants in the study according to inclusion and exclusion criteria (Supplementary Methods S1). The HN5000 cohort was screened to identify patients alive and disease free 12 months after diagnosis. The percentages refer to the percentage of total screened.

Baseline characteristics are presented in Table 1. Deceased subjects were more often male, older, with higher cancer stages, more comorbidities, had more HPV negative tumours, had lower income, less education, smoked more and consumed more alcohol than patients alive 3 years after diagnosis. Of the patients alive 3 years after diagnosis, subjects missing QoL were more often male, had more comorbidities, lower household income, were not married, had lower education, lived in more deprived areas and smoked more compared to those with measured QoL (Supplementary Table S5).

**Table 1.**
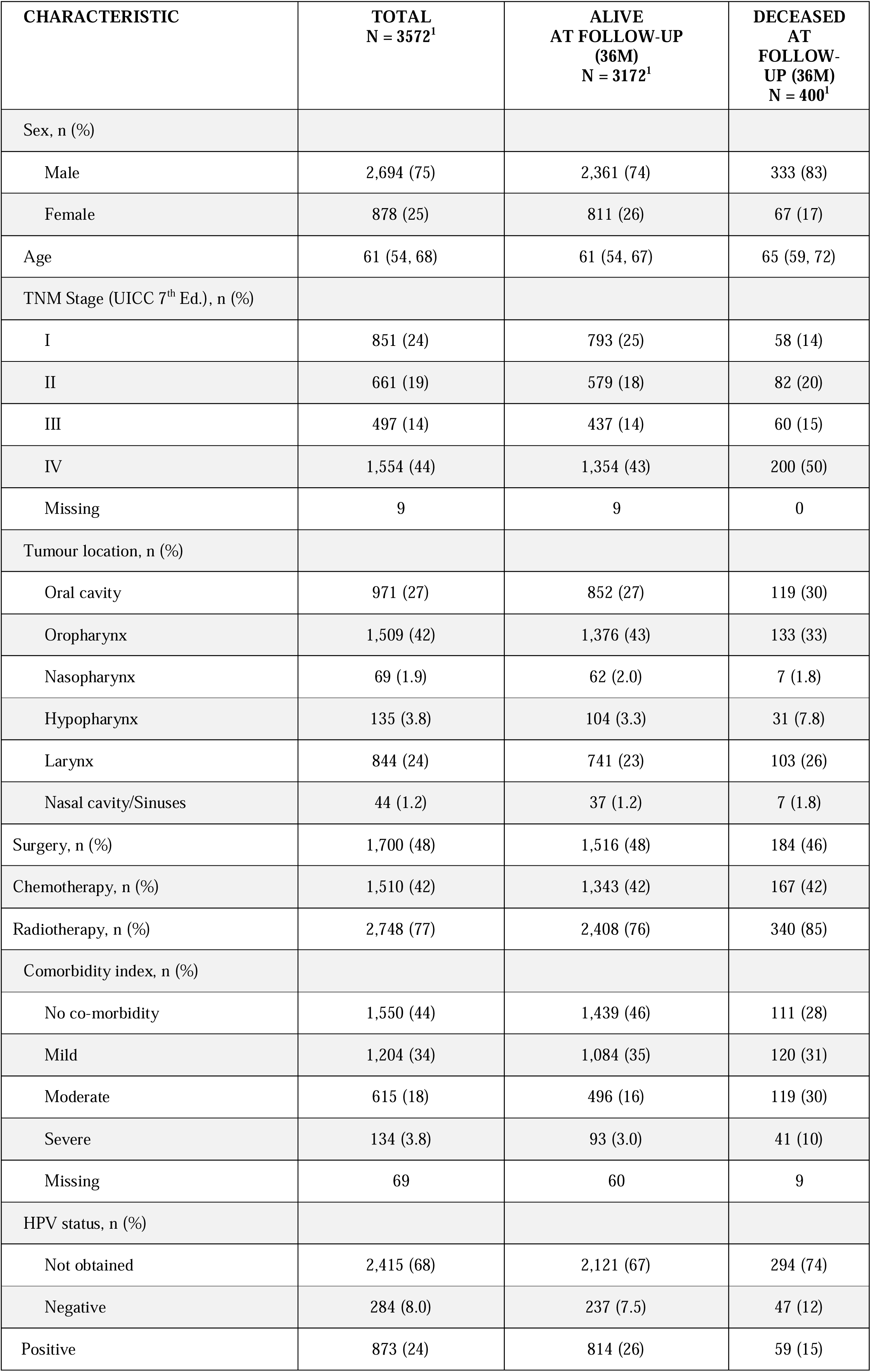
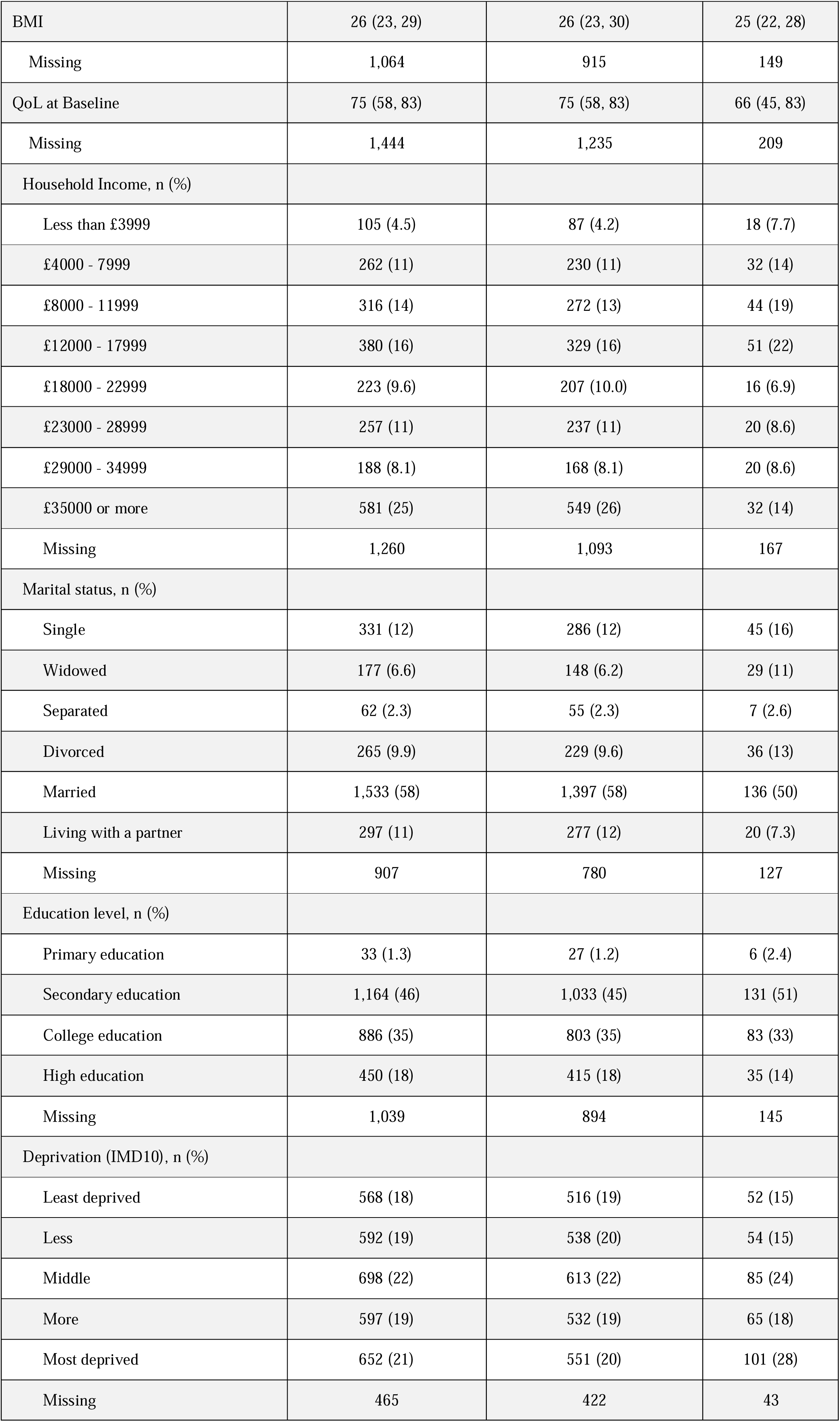
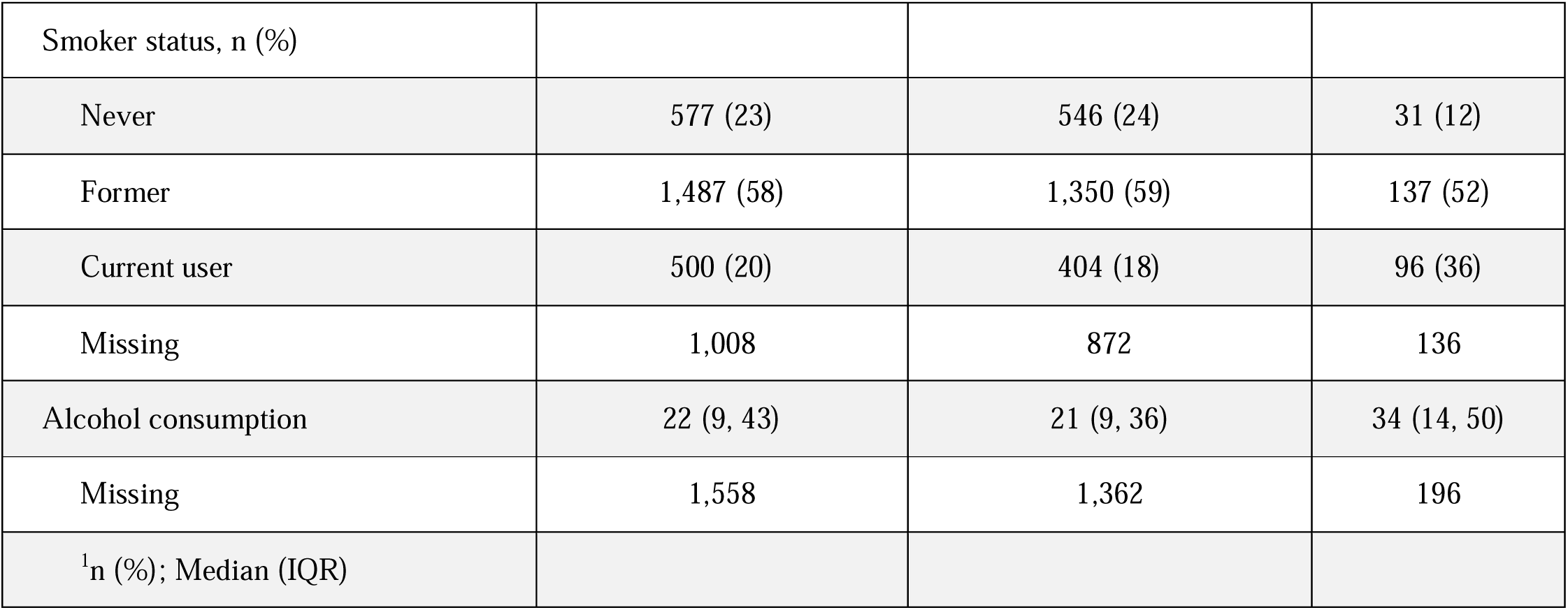
Descriptive statistics for the included cohort and stratified by vital status at prediction time (36 months follow-up from diagnosis). Median and interquartile are shown for continuous characteristics. Counts and percentages are shown for the categorical characteristics.

| CHARACTERISTIC | TOTAL<br>N = 3572<sup>1</sup> | ALIVE<br>AT FOLLOW-UP<br>(36M)<br>N = 3172<sup>1</sup> | DECEASED<br>AT<br>FOLLOW-UP (36M)<br>N = 400<sup>1</sup> |
| --- | --- | --- | --- |
| Sex, n (%) |  |  |  |
| Male | 2,694 (75) | 2,361 (74) | 333 (83) |
| Female | 878 (25) | 811 (26) | 67 (17) |
| Age | 61 (54, 68) | 61 (54, 67) | 65 (59, 72) |
| TNM Stage (UICC 7<sup>th</sup> Ed.), n (%) |  |  |  |
| I | 851 (24) | 793 (25) | 58 (14) |
| II | 661 (19) | 579 (18) | 82 (20) |
| III | 497 (14) | 437 (14) | 60 (15) |
| IV | 1,554 (44) | 1,354 (43) | 200 (50) |
| Missing | 9 | 9 | 0 |
| Tumour location, n (%) |  |  |  |
| Oral cavity | 971 (27) | 852 (27) | 119 (30) |
| Oropharynx | 1,509 (42) | 1,376 (43) | 133 (33) |
| Nasopharynx | 69 (1.9) | 62 (2.0) | 7 (1.8) |
| Hypopharynx | 135 (3.8) | 104 (3.3) | 31 (7.8) |
| Larynx | 844 (24) | 741 (23) | 103 (26) |
| Nasal cavity/Sinuses | 44 (1.2) | 37 (1.2) | 7 (1.8) |
| Surgery, n (%) | 1,700 (48) | 1,516 (48) | 184 (46) |
| Chemotherapy, n (%) | 1,510 (42) | 1,343 (42) | 167 (42) |
| Radiotherapy, n (%) | 2,748 (77) | 2,408 (76) | 340 (85) |
| Comorbidity index, n (%) |  |  |  |
| No co-morbidity | 1,550 (44) | 1,439 (46) | 111 (28) |
| Mild | 1,204 (34) | 1,084 (35) | 120 (31) |
| Moderate | 615 (18) | 496 (16) | 119 (30) |
| Severe | 134 (3.8) | 93 (3.0) | 41 (10) |
| Missing | 69 | 60 | 9 |
| HPV status, n (%) |  |  |  |
| Not obtained | 2,415 (68) | 2,121 (67) | 294 (74) |
| Negative | 284 (8.0) | 237 (7.5) | 47 (12) |
| Positive | 873 (24) | 814 (26) | 59 (15) |
| BMI | 26 (23, 29) | 26 (23, 30) | 25 (22, 28) |
| Missing | 1,064 | 915 | 149 |
| QoL at Baseline | 75 (58, 83) | 75 (58, 83) | 66 (45, 83) |
| Missing | 1,444 | 1,235 | 209 |
| Household Income, n (%) |  |  |  |
| Less than £3999 | 105 (4.5) | 87 (4.2) | 18 (7.7) |
| £4000 - 7999 | 262 (11) | 230 (11) | 32 (14) |
| £8000 - 11999 | 316 (14) | 272 (13) | 44 (19) |
| £12000 - 17999 | 380 (16) | 329 (16) | 51 (22) |
| £18000 - 22999 | 223 (9.6) | 207 (10.0) | 16 (6.9) |
| £23000 - 28999 | 257 (11) | 237 (11) | 20 (8.6) |
| £29000 - 34999 | 188 (8.1) | 168 (8.1) | 20 (8.6) |
| £35000 or more | 581 (25) | 549 (26) | 32 (14) |
| Missing | 1,260 | 1,093 | 167 |
| Marital status, n (%) |  |  |  |
| Single | 331 (12) | 286 (12) | 45 (16) |
| Widowed | 177 (6.6) | 148 (6.2) | 29 (11) |
| Separated | 62 (2.3) | 55 (2.3) | 7 (2.6) |
| Divorced | 265 (9.9) | 229 (9.6) | 36 (13) |
| Married | 1,533 (58) | 1,397 (58) | 136 (50) |
| Living with a partner | 297 (11) | 277 (12) | 20 (7.3) |
| Missing | 907 | 780 | 127 |
| Education level, n (%) |  |  |  |
| Primary education | 33 (1.3) | 27 (1.2) | 6 (2.4) |
| Secondary education | 1,164 (46) | 1,033 (45) | 131 (51) |
| College education | 886 (35) | 803 (35) | 83 (33) |
| High education | 450 (18) | 415 (18) | 35 (14) |
| Missing | 1,039 | 894 | 145 |
| Deprivation (IMD10), n (%) |  |  |  |
| Least deprived | 568 (18) | 516 (19) | 52 (15) |
| Less | 592 (19) | 538 (20) | 54 (15) |
| Middle | 698 (22) | 613 (22) | 85 (24) |
| More | 597 (19) | 532 (19) | 65 (18) |
| Most deprived | 652 (21) | 551 (20) | 101 (28) |
| Missing | 465 | 422 | 43 |
| Smoker status, n (%) |  |  |  |
| Never | 577 (23) | 546 (24) | 31 (12) |
| Former | 1,487 (58) | 1,350 (59) | 137 (52) |
| Current user | 500 (20) | 404 (18) | 96 (36) |
| Missing | 1,008 | 872 | 136 |
| Alcohol consumption | 22 (9, 43) | 21 (9, 36) | 34 (14, 50) |
| Missing | 1,558 | 1,362 | 196 |
| <sup>1</sup>n (%); Median (IQR) |  |  |  |

### Summary of development

We developed conformal LASSO and XGBoost models for the continuous QoL outcome (n=1662), using either the limited or extended set of predictors, and estimated the predictive distribution. For predicting mortality at three years after diagnosis (no censoring), we used logistic regression with L1-penalty, with n=3572 subjects and 400 events. The conformal LASSO and XGBoost models were used to predict the probability of QoL decline (Eq.5), with 238 QoL declines in 1351 subjects. The combined models for predicting QoL decline or death were then applied to 1751 observations and 638 events (Supplementary Table S6).

### Model specification

The full list of estimated parameters are given in Tables S7, S8 and S9. To use our prediction model, one can either directly interface with our API (https://bd4predict.azurewebsites.net) or employ our dashboard – the BD4QoLPredict tool (https://ocbe-uio.github.io/bd4predict). Users input demographic, clinical, and QoL data, and the model predicts the joint probability of survival and QoL decline, estimated QoL score with prediction intervals, and the predictive conformal distribution, imputing missing. Imputed features are displayed in the dashboard for further analysis (Supplementary Fig. S2). Complete API documentation and dashboard guidelines are provided in the respective repositories.

### Performance evaluation

Table 2 presents the performance metrics for the joint models and submodels developed with LASSO and XGBoost. Overall, models developed with the limited set of predictors present similar discrimination and better calibration compared with the extensive counterpart. All models had a moderate degree of miscalibration in different directions. Considering the usability and interpretability of simpler models, the close performance and better calibration, LASSO/limited is our top deployment choice. We evaluated the LASSO/limited model in two groups of subjects that present high and low propensity scores for missing outcome and found similar performance (Supplementary Table S10).

**Table 2.**
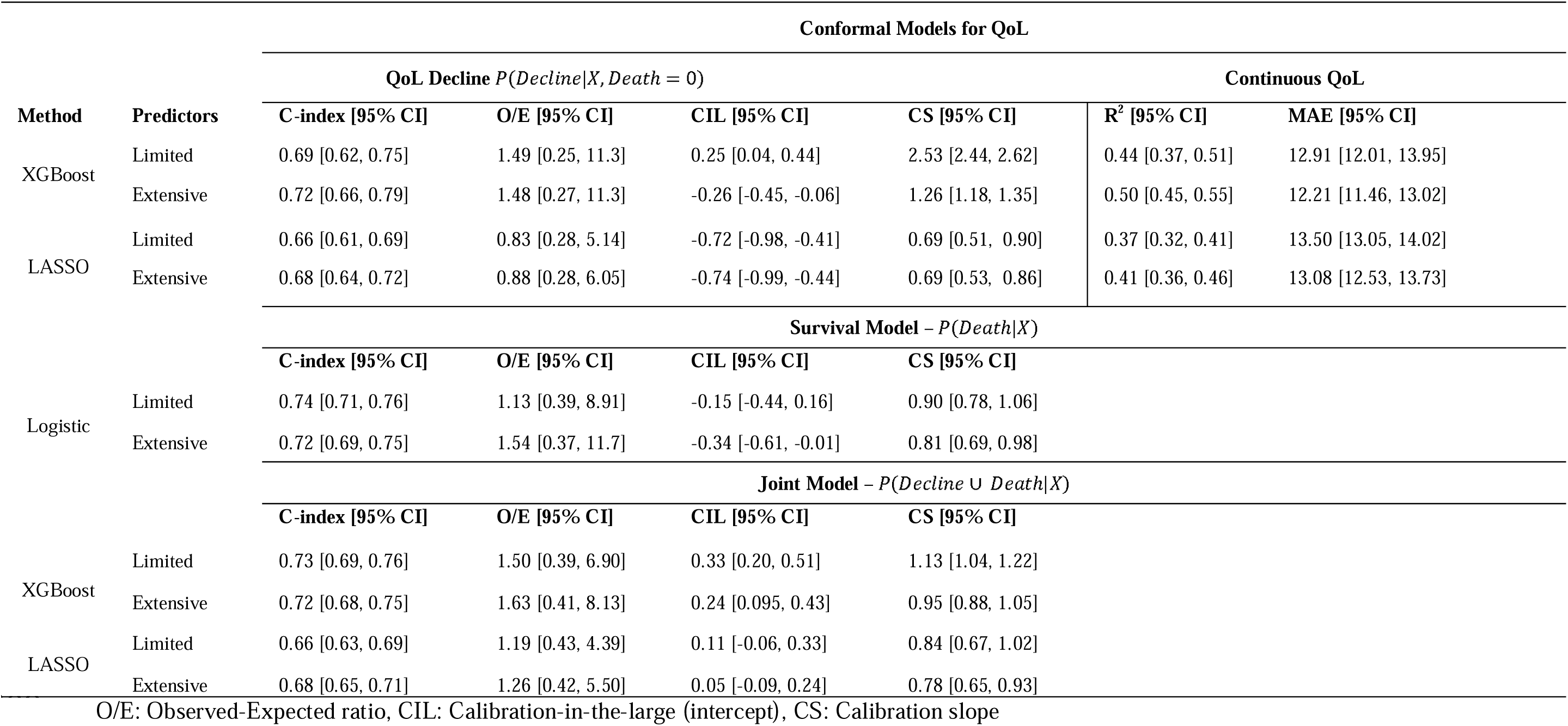
Discrimination and continuous QoL score prediction performance were assessed with bootstrap 666 .638+ estimator and calibration estimates were obtained with the .638 estimator. Models were assessed with two different sets of predictors (limited and extensive).

Figure 2 presents the calibration plots and histograms for (a) continuous QoL conditional on survival, (b) for the probability of “QoL decline or death” (joint model), (c) conditional QoL decline, and (d) probability of being deceased at 36 months after diagnosis. The models present skewed prediction distributions, in agreement with the distributions observed in the development data. Moderate miscalibration occurs at continuous outcome extremes and in the conditional QoL decline model with systematically risk overestimation. Weak miscalibration is observed for the joint model with risk underestimation probabilities ranging from 0.3 to 0.4. The survival model showed excellent calibration. Overall, the models are reasonably well calibrated with good alignment with the observed data.

**Figure 2.**
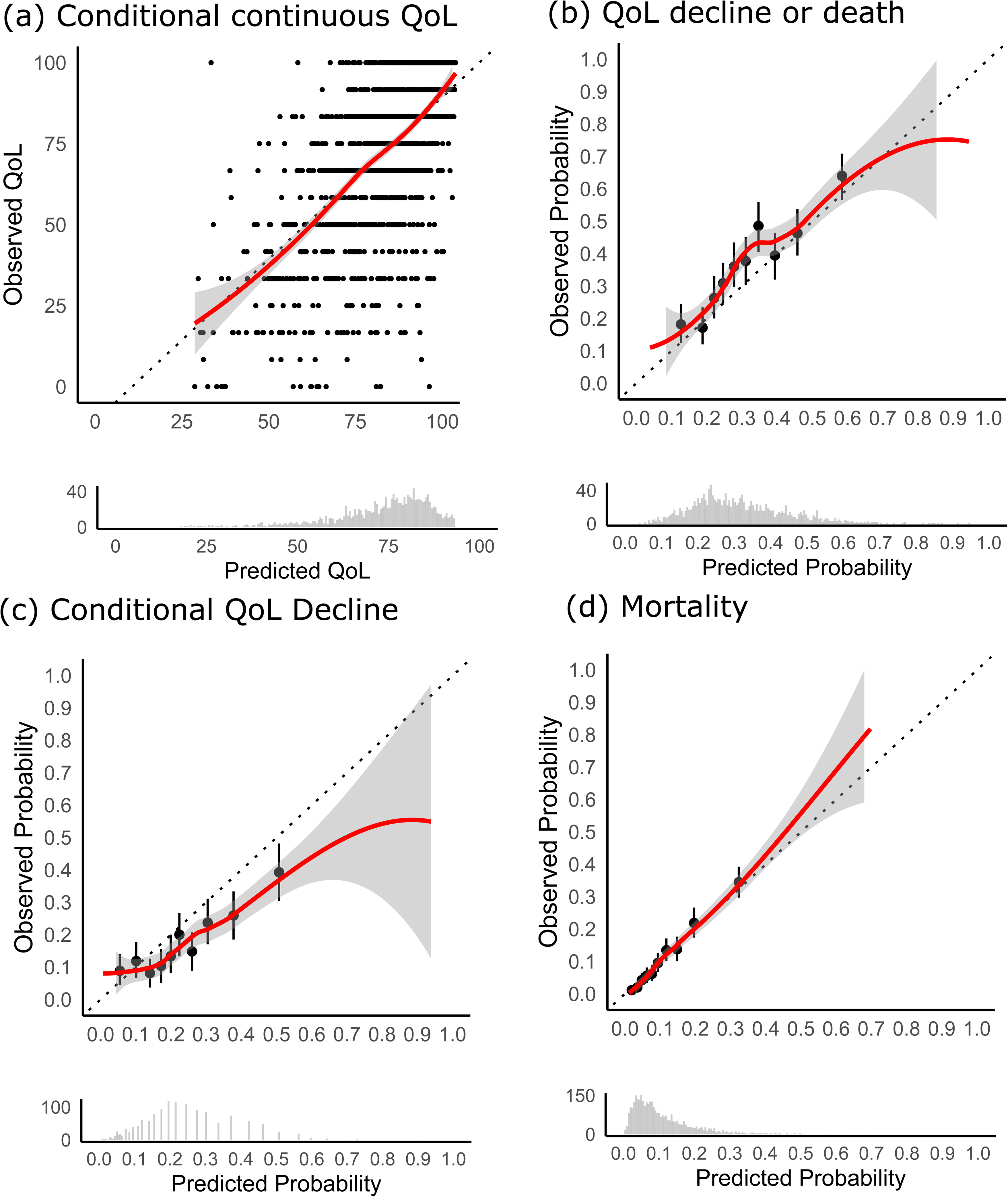
Calibration plots for the predictive models at three years after diagnosis. (a) Continuous outcome QoL. (b) Probability of decline or death. (c) Conditional probability of QoL decline. (d) Probability of survival. In each plot, the x-axis represents the predicted values, while the y-axis represents the observed values. The black dots represent the estimated observed probabilities for different bins of predicted probabilities and 95% confidence intervals. The solid red lines are smoothed curves obtained with LOESS, and the grey lines denote the 95% confidence intervals. The 45-degree diagonal lines are the reference for perfect calibration. At the bottom of each calibration the predicted probability histogram is shown.

In external validation LASSO- and XGboost-limited performed similar to the internal validation performance for the continuous QoL with MAEs of 12.97 and 12.28, and R2 of 0.353 and 0.414, respectively. Discriminative performance was higher than the one observed in internal validation with C-statistics of 0.799 and 0.795 for LASSO and XGBoost respectively. The models presented overall good calibration in the external data but with large confidence intervals, with slight indication of systematic risk of underestimation for LASSO and overestimation for XGBoost (Figure 3). The survival submodel was validated with an AUC of 0.60, and calibration metrics E/O = 0.64, calibration in the large = 0.15 and calibration slope 0.64. The calibration curve (Figure S3) showed an underprediction of mortality, particularly close to the mean predicted mortality (approximately 0.1).

**Figure 3.**
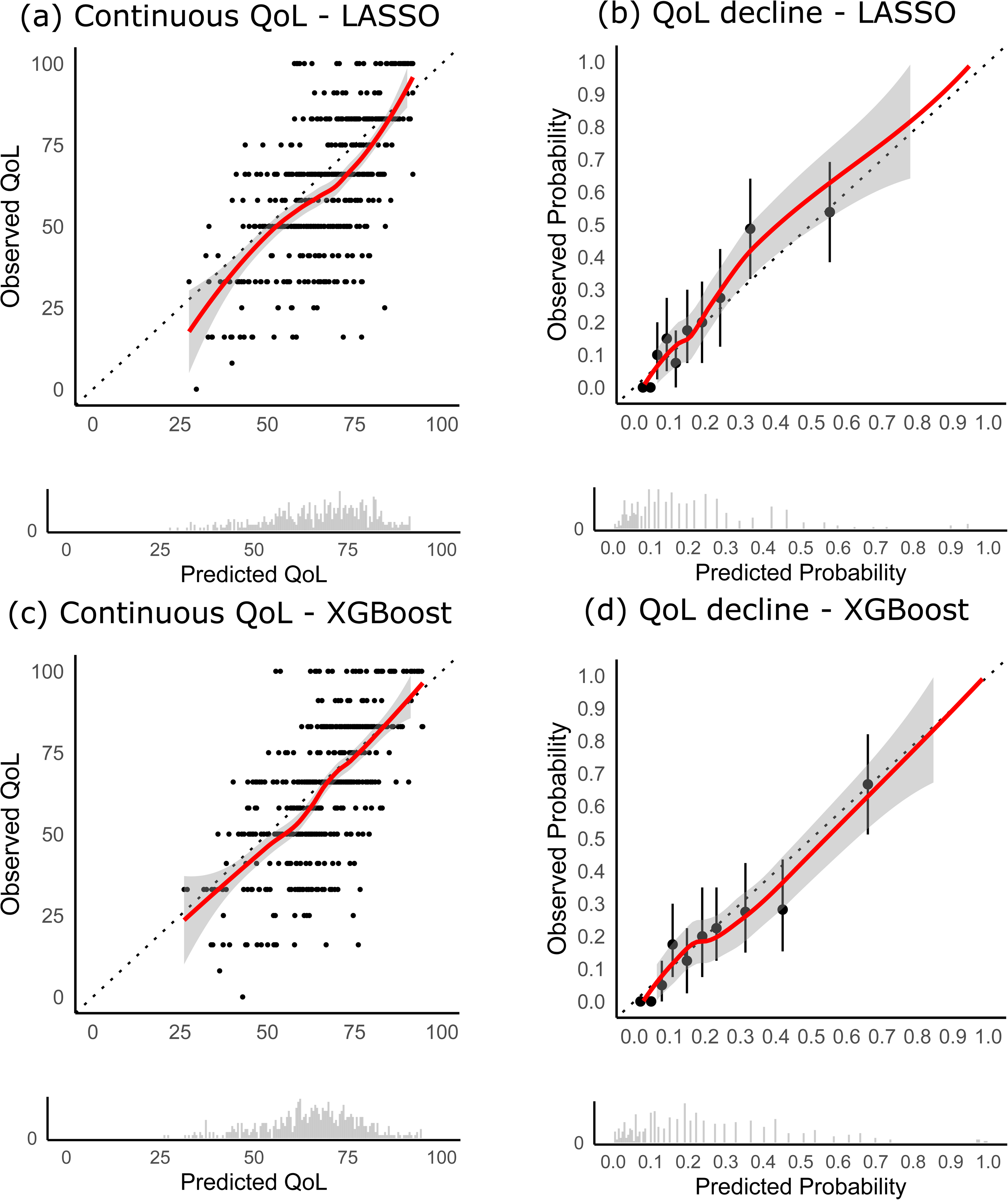
External validation for models developed with the limited set of predictors. Figures (a) and (b) depict calibration curves for the continuous QoL and conditional QoL decline for LASSO, where (c) and (b) show calibration for XGBoost. The histograms show distributions of predicted values. The black dots represent the estimated observed probabilities for different bins of predicted probabilities and 95% confidence intervals. The solid red lines are smoothed curves obtained with LOESS, and the grey lines denote the 95% confidence intervals. The 45-degree diagonal lines are the reference for perfect calibration. At the bottom of each calibration the predicted probability histogram is shown.

### Model Explainability

SHAP values quantify the effect of the covariates on the prediction for each patient. Figure 4 collects the SHAP values for all individuals for (a) mortality, conditional QoL with (b) LASSO, (c) XGBoost, and (d) the SHAP interactions from XGBoost. Common predictors of mortality, such as age and TNM stage, are associated with increased probability of death. Conditional QoL is mainly linked with QoL measured at earlier time points suggesting strong autocorrelation. The strongest interactions are observed between QoL at 12 months after diagnosis and other functional scales, household income, BMI, alcohol consumption, and age.

**Figure 4.**
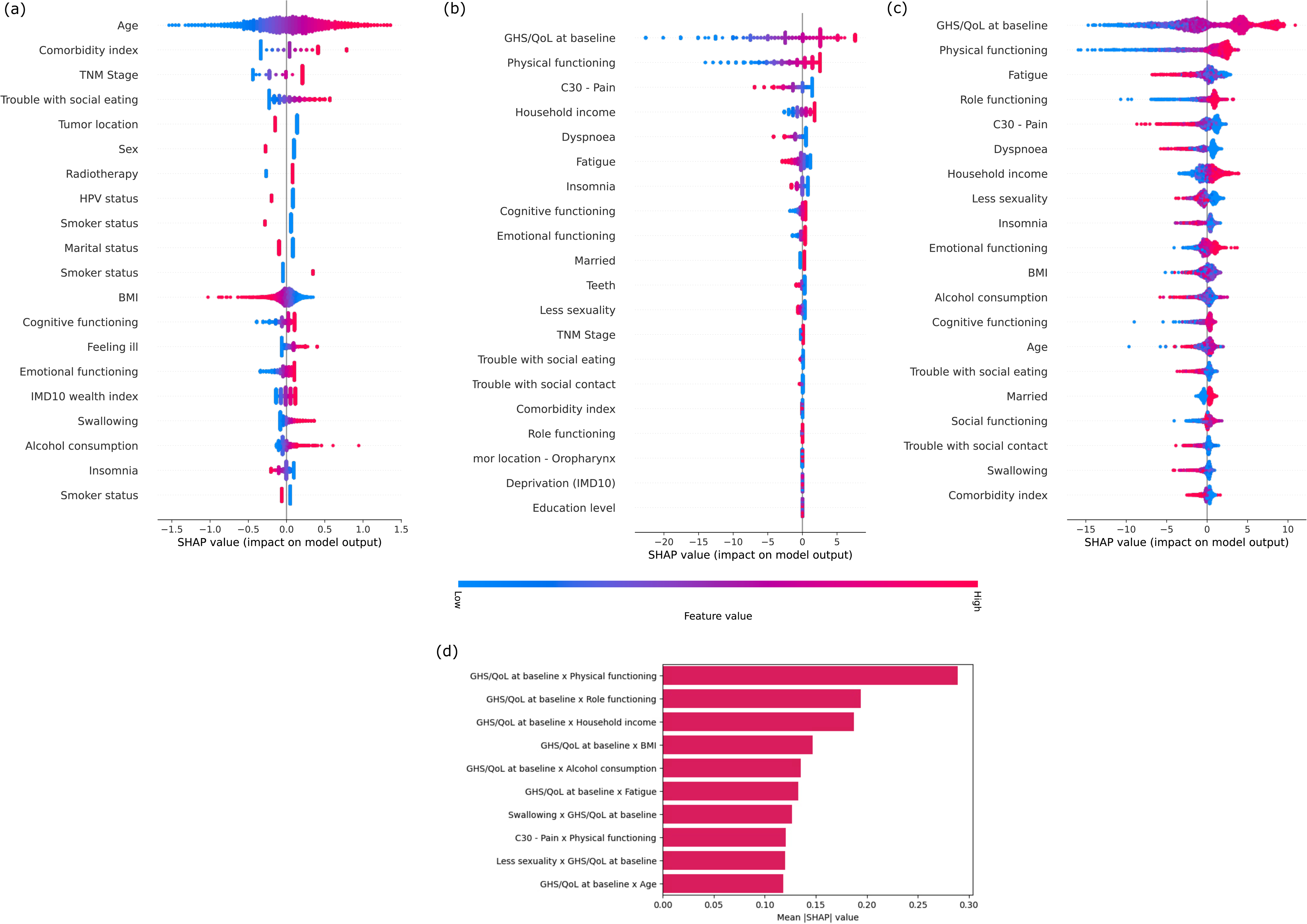
SHAP values associated with prediction models for (a) mortality, conditional QoL with (b) LASSO, (c) XGBoost, and (d) the respective top 1-percentile of mean absolute SHAP interaction values.

## Discussion

### Summary of main findings

This study introduces a joint probability approach for prognostic models for conditional outcomes, that can be tailored to answer relevant probabilistic questions at inference time. We applied this framework and developed a prognostic model for QoL in a large cohort of HNC patients, which provided a robust dataset for model development. The results indicate that the joint model, which combines predictions of survival and QoL, offers a more comprehensive prognostic picture compared to models that only consider the conditional outcome.

We developed and made available the BD4QoLPredict API and dashboard, to facilitate access to the models either programmatically or via a user-friendly interface. This sets a high standard as one of the first ready-to-use prognostic tools developed following rigorous methodology to minimize bias and to provide probabilistic QoL predictions for HNC patients. The vast majority of previous studies were aimed at prognostic factor discovery in this population(22,29), but no previous prognostic tool is available.

The joint probability model has significant potential for clinical use. By providing predictions for both survival and QoL decline together, the model can help clinicians identify high-risk patients who may benefit from early interventions aimed at improving both survival and quality of life. This approach can also inform healthcare resource planning, ensuring that efforts are directed towards patients with the greatest need.

External validation showed good discriminative performance. However, future research should focus on external validation of the model in different populations and settings to confirm generalizability.

### Strengths and limitations

A key strength of the joint modelling approach for conditional outcomes is that one maintains a probabilistic interpretation of the predictions. Compare this to commonly applied alternatives in prediction model development (i.e., model development in the subset of individuals where the conditional outcome is defined or imputation of the conditional outcome for those where the conditional outcome is not defined(30)). The former results in conditional probabilities or classifications, but this may be not explicitly acknowledged and thus result in invalid interpretations. The latter makes interpretation of the predictions difficult as values are imputed for individuals where the conditional outcome is not defined.

Another key strength of the joint modelling approach is that it results in a flexible model that, when coupled with an API and dashboard, can be tailored to the relevant questions in the field. One can ask questions conditional on the post-baseline event occurring or about the joint probability of the post-baseline event occurring and about some state/range for the conditional outcome.

A limitation of the joint modelling approach could be in datasets where the relevant post-baseline event is not observed and where it is impossible to distinguish between outcomes missing due to being conditionally defined or missing due to other causes.

An important feature of the joint modelling approach is that it is probabilistic, which provides direct interpretation for the predictions in contrast with methods that provide only score-based binary classification. We can predict the continuous measurement with prediction intervals, the probability of the outcome laying in a post-defined range, and the probability of the outcome not being observed.

One key strength in our application is the use of a large, well-characterized cohort of HNC patients. The study also employed advanced statistical techniques, including conformal prediction and SHAP, to ensure robust model development and interpretation. We followed the highest methodological standards for model development and performance assessment with internal and external validation, by estimating important metrics such as calibration and employing bias-corrected bootstrap estimators.

However, there are some limitations to consider. The sample size, while substantial, may still pose a risk of overfitting, particularly for the extended set of predictors. The study also faced challenges with missing data, but these were addressed using imputation methods in the model development and propensity scores for model evaluation. In addition, future research can explore different optimization strategies for the joint model estimation by combining the objective functions of the submodels rather than estimating them separately.

The modest AUC performance observed suggests that while the model has predictive capacity, it should be used as a supplementary tool alongside clinical judgment. The model’s ability to predict both survival and QoL provides a more holistic view of patient prognosis, unlike a conditional approach that assumes survival. This comprehensive perspective is crucial for clinical decision-making and resource allocation.

## Conclusion

In conclusion, this study presents a novel joint probability approach to prediction modelling for conditional outcomes, with application in HNC patients. We present a comprehensive prognostic tool that can aid in clinical decision-making and resource allocation. We underscore the importance of recognising conditional outcomes when developing prognostic prediction models.

## Declarations

### Data Availability

Non-sensitive data generated during model development is shared in this manuscript and the respective supplementary material. The BD4QoL historical cohort dataset is hosted by the Services for Sensitive Data (TSD) at the University of Oslo. Access to the data may be granted by the data owners upon application. The data that support the findings of this study are available from head and neck 5000. Further information may be found on the Head and Neck 5000 website: https://www.headandneck5000.org.uk/information-for-researchers.

### Ethical Approval

The study protocol received ethical approval by the Norwegian Regional Committee for Medical and Health Research Ethics (REK) South-East D under application number 154191. The data is stored in compliance with GDPR legislation in the secure server for sensitive data at the University of Oslo (TSD/USIT) and access is granted to authorised collaborators included in the ethical approval. The HN5000 study was approved by the National Research Ethics Committee (South West Frenchay Ethics Committee, reference number 10/H0107/57, 5 November 2010) and the Research and Development departments of participating NHS Trusts. Informed consent was obtained from all patients recruited to HN5000. BD2Decide study was approved by institutional research ethics board with identifier N. INT 65/16. UMM1 and UMM2 received ethical approval from the Leipzig University Ethics Committee. UMM3 was approved by Landesärztekammer Rheinland-Pfalz ethics committee under approval number 367 837.281.14 (9520).

## Acknowledgements

This work was performed on the TSD (Tjeneste for Sensitive Data) facilities, owned by the University of Oslo, operated and developed by the TSD service group at the University of Oslo, IT-Department (USIT). The statistical analysis and data harmonisation were performed on resources provided by Sigma2 - the National Infrastructure for High Performance Computing and Data Storage in Norway. The Authors thank the researchers and clinicians who designed the HN5000 study, the research, laboratory and clinical staff who supported the conduct of the study; and the people with head and neck cancer who took part.

## Contribution statement

ML, AF, MM-S contributed to the conceptualisation. ML, AF, MM-S, AB-J, AA, EIFF contributed to the methodology. ML, AF contributed to the supervision. ML, MM-S drafted the first draft of the manuscript. SS, LL, HN5000 executive group and study pathologist contributed data. MM-S, EIFF contributed to the data analysis. ML, MM-S, EIFF, AB-J, AA, LL-P, IA, HN5000 executive group had full access to all study data. Access for all authors to the raw data was not feasible due to legal restrictions. SS and KT provided expertise on quality of life measurement. LL-P, IA, AB-J, MFC-U, G-F and AA provided expertise on the evaluation of machine learning models. All authors had access to the data presented in this study. All authors reviewed and approved the manuscript.

## Declaration of generative AI and AI-assisted technologies in the writing process

During the preparation of this work the authors used GPT UiO in order to reduce the word count in a few sentences. After using this tool/service, the authors reviewed and edited the content as needed and take full responsibility for the content of the publication.

