## Supplemental Information for "Joint probability approach for prognostic prediction of conditional outcomes: application to quality of life in head and neck cancer survivors"

* - Author whom correspondence should be addressed.

### Supplementary Methods

#### S1. Eligibility criteria for inclusion in the model development

##### Inclusion criteria

1. Non-metastatic head and neck cancers from one of the following subsites (ICD in annex 1): oral cavity, hypopharynx, larynx, oropharynx, nasopharynx, major or minor salivary glands, nasal cavity, paranasal sinuses.

2. Having received and concluded treatments with curative intent at time of study inclusion.

3. Being alive and disease-free at last post-treatment follow-up.

4. Stage I, II, III, IVa or IVb according to TNM 7th edition (1).

5. Age ≥18 years.

#### Exclusion criteria

1. Histology other than squamous cell carcinoma and salivary gland carcinomas Thyroid cancers, neuroendocrine tumours and non-epithelial HNC (e.g., melanoma, sarcoma, etc.) are excluded.

2. Distant metastases at the time of study entry.

3. Any previous HNC unrelated to the primary HNC for which the participant was treated; premalignant lesions (e.g., leukoplakia, erythroplakia, lichen etc.) are allowed.

4. Subjects with previous malignancies (except localized non-melanoma skin cancers, and the following in situ cancers: bladder, gastric, colon, esophageal endometrial, cervical/dysplasia, melanoma, or breast) unless a complete remission was achieved at least 5 years prior to study entry and no additional therapy is required during the study period.

### S2. Adverse event probability derivation

We can answer different prognostic clinical questions by rewriting slightly Eq. (5) from the main text. For example, the probability of any adverse events, having a QoL decline or dying, can be obtained by using the complement of survival $D = S^c$,

$$P\left(QoL \le QoL_{t_0} - 10 \cup D \middle| X\right) = P\left(QoL \le QoL_{t_0} - 10,\ S \middle| X\right) + P(D|X). \quad \text{(S1)}$$

Replacing Eq. (6) in (5)

$$P\left(QoL \le QoL_{t_0} - 10 \cup D \middle| X\right) = P\left(QoL \le QoL_{t_0} - 10 \middle| X, S\right) P(S|X) + P(D|X), \quad \text{(S2)}$$

and re-writing it in terms of adverse events only for consistency

$$P\left(QoL \le QoL_{t_0} - 10 \cup D \middle| X\right) = P\left(QoL \le QoL_{t_0} - 10 \middle| X, D = 0\right)\left(1 - P(D|X)\right) + P(D|X). \quad \text{(S3)}$$

### S3. Minimum sample size estimation

Considering the minimum $R^2$ of 0.30 based on a univariate model with QoL at baseline as predictor, shrinkage coefficient of 0.90, outcome standard deviation of 22.19 and mean 73.90, the estimated sample size required is 1430 samples for 59 candidate predictor parameters (see Table S2). For the extended set of 171 candidate predictor parameters the required sample 4249 observations (see Table S3). We are aware that variable selection with least absolute shrinkage and selection operator (LASSO) can also be affected by small sample size (2), but the development of models with both sets of predictors is used as sensitivity analysis.

### S4. Propensity score approach

We developed a propensity score model to estimate the probability of missing the outcome measurement. We constructed the propensity scores by estimating a logistic regression model with a concise set of variables that have demonstrated associations with missing data in survey research, namely: sex, age, household income, marital status, educational attainment, and a regional deprivation index. Additionally, recognizing that overall health may also affect survey participation, we incorporated several health-related variables: smoking status, alcohol consumption, and scores from the QLQ-C30 questionnaire assessing fatigue, pain, and QoL/GHS. Furthermore, we included variables for received treatment, comorbidity index, and TNM stage, acknowledging their potential influence on response rates. We used the propensity scores to evaluate model performance on subjects that presented similar characteristics to the group with missing outcomes based on inverse probability weighting.

### S5. Model development

The continuous QoL score was modelled with multivariable probabilistic models using conformity scores (3), which provides conformal predictive distributions and quantifies both the variability from the new prediction and the uncertainty from estimating the unknown parameters (4). We used MAPIE to estimate the conformal models, which combines the conformity scores (residuals) of an ensemble of regression functions and the prediction uncertainty from cross-validation, where any machine learning method can be used as the predictor of a non-parametric predictive distribution. However, other approaches can be applied such as Bayesian estimation. We employed a conformal method which uses the CV+ estimator (5) with 5-fold cross validation to estimate the prediction uncertainty. The predicted risk of QoL decline is obtained from the predicted distribution. The base estimators for the probabilistic models were linear regression with LASSO and boosted trees with the XGBoost implementation. The predicted risk of death was modelled with logistic regression and LASSO penalty. Model hyperparameter tuning was performed with Bayesian optimisation (6) by minimising the average of 5-fold cross validation of the mean squared error (MSE) for the continuous outcome, and the C-statistics for the logistic model. No method was employed for handling class unbalance, considering that the main goal was to estimate accurate predicted probabilities.

### S6. Internal validation

Internal validation was performed with bootstrap using 200 resamples with replacements. We applied the .632+ estimator to obtain the discriminative performance estimates, which correct for overly pessimistic performance in the out-of-bag samples and the overly optimistic evaluation from assessing the model in the development data (7). This estimator requires the definition of a non-informative performance or the performance of a benchmark model. For the absolute mean error (MAE), and the unadjusted coefficient of determination $R^2$, we determined the non-information performances for a model that always predicts the population median value. For C-statistics we used 0.5 as non-informative baseline performance. For assessing calibration, we applied the .638 bootstrap estimator (8) to obtain the observed-expected ratio (O/E), calibration-in-the-large (CIL), and calibration slope (CS). Calibration plots with flexible curves were obtained using locally estimated scatterplot smoothing (LOESS). The dataset exhibits a substantial quantity of missing outcomes, which can hinder the process of internal validation. To address this issue, we assessed model performance using a propensity score approach designed to estimate the probability of missing outcomes.

### S7. The Head and Neck 5000 study

HN5000 is a large UK-based study of individuals with head and neck cancer. It is sponsored by University Hospitals Bristol and Weston NHS Foundation Trust (UHBW) and conducted in collaboration with the University of Bristol (9,10). From 2011 to 2014, 5,511 participants were recruited across 76 UK centres, making it one of the world's largest prospective cohort studies in this field. Over 200 variables were collected from diagnosis with up to three years of follow-up. Information includes clinical, demographic, quality of life assessments, and physical and mental health data. The cohort has been followed up to the present day. Inclusion criteria were age 16 or older with a new diagnosis of primary head and neck cancer or cancer of unknown primary (CUP). Exclusion criteria included people with reduced mental capacity as per legal definitions, recurrent or a second head and neck cancer, skin cancer, lymphomas, and a histological diagnosis of Carcinoma in Situ (unless upstaged to invasive disease following imaging review/MDT discussion. Patients were enrolled at the time of diagnosis and before definitive treatment commenced unless the diagnostic biopsy procedure formed part of the surgical management Comprehensive information can be sourced viattps://headandneck5000.org.uk.

### S8. External validation datasets for the quality of life submodel

External validation for the conformal model was performed with independent data from the University of Mainz (Germany) and the Istituto Nazionale dei Tumori (Italy). The data contains N=497 subjects with measured QoL and meeting eligibility criteria in this study. From those, N=398 had the QoL decline also measured, and N=79 (20%) presented a QoL decline. We evaluated the continuous outcome with R-squared, MAE, and discriminative performance (C-statistics) and overall calibration with LOESS curves for the conditional QoL decline. Missing data in the predictors were imputed with KNN imputer built in the development data. For validating the survival submodel we used a larger subset of the BD2Decide data that contains vital status of 281 patients, with 26% events (N=73)

The external validation data is composed by data from 4 different studies. The UMM1(11) was a prospective cohort study in patients before and after total laryngectomy (TLE). Further eligibility criteria were written informed consent and age of 18 years or older. The patients were interviewed in a face-to-face setting before the surgery (t1), shortly before discharge from the hospital (t2), at the end of rehabilitation (t3), one year after baseline (t4) as well as two (t5) and three years (t6) after baseline. Participants also completed self-administered questionnaires, including the EORTC QLQ-C30 and the EORTC QLQ-H&N35. A total of 389 patients were enrolled between the years 2001-2011 from 13 hospitals in Germany. The UMM2 data (12) comes from a similar study, but this time in patients who were scheduled for partial laryngectomy (PLE). The study design and data collection were in parallel with the UMM1 study up to t4. Data collection began in 2007 and ended in 2015. A sample of 391 patients were enrolled from 16 hospitals in Germany.

The UMM3 data (13) comes from an international validation study for the update of the EORTC QLQ-H&N35 questionnaire, the HN43 Phase IV study. In total, 812 patients from 18 countries in Europe, the Americas and Asia were enrolled. Patients with cancer of the larynx (ICD-10 code C32), lip (C00), oral cavity (C01-06), salivary glands (C07-08), oro-hypopharynx (C09-10, C12-14), nasopharynx (C11), nasal cavity (C30), nasal sinuses (C31), sarcoma in the head and neck region (C49), and lymph node metastases from Missing primary in the head and neck area (C77, C80.0).were included. There were no restrictions regarding stage, recurrence status, or treatments planned or performed. Patients with a tumour of the eyes, orbit, thyroid, skin (even if in the head and neck area), or lymphomas in the head and neck region were excluded. Patients completed the questionnaires up to 14 days before start of treatment (t1), three months (t2), and six months thereafter (t3).

The Big Data to Decide (BD2Decide) project was a European multicentre observation study including 1537 HNC patients from Italy, Germany and the Netherlands(14). BD2Decide was approved by the Ethical Committee of the Fondazione IRCCS Istituto Nazionale dei Tumori (Milan, Italy) in 2016 and it has two identifiers: INT65-16 and INT66-16. BD2Decide received funding from the European Union Horizon 2020 Framework Programme, Grant/Award Number: 689715. The aim of this project was to develop a multisource database to allow for prognostic prediction modelling in loco-regionally advanced HNC patients. The database was made of two cohorts: retrospective (diagnosis 2008-2014), and prospective (diagnosis 2015-2017). Main inclusion criteria were diagnosis of head and neck squamous cell carcinoma (HNSCC); stage III and IVA/B (based on AJCC/UICC seventh edition); receiving treatments with curative intent; availability of pre-treatment tumour specimen for biological analysis; availability of pre-treatment imaging scans for radiomic analysis; for patients enrolled prospectively, PROs were collected (EORTC C30, EORTC HN35 and EQ-5D-5L). The BD4QoL study included the prospective BD2Decide patients enrolled at one of the Italian cancer centres.

For external validation we filtered only patients that presented the EORTC QLQ-C30 measurement at the follow-up, which reduced the dataset to 497 observations. All patients included complied with the eligibility criteria used to select the population for model development. Basic characteristic can be found in Table S11. Survival data was not recorded, so this data was used only for the quality of life submodel validation.

### S9. External validation for the survival submodel

We used BD2Decide data that contained information on survival (see Table S12 for characteristics) for external validation (see Supplementary Methods S8 for description of the study). This dataset was particularly suited for external validation because it contained many of the variables that were used in the development data, including the comorbidity index.

To align external survival data with the development data, patients that died or were censored prior to 1 year after diagnosis were excluded, leaving a total of 384 patients remaining. Of these, 27% were censored between 1 year and 3 years after survival, and were therefore removed from the external validation, leaving a total of 281 eligible patients. The overall observed mortality between 1 and 3 years since diagnosis was 26%.

Coefficients from the logistic survival submodel were used to predict mortality 3 years after diagnosis (2 years after the baseline) in the external data. Variables in the external data were harmonized to fit the same format as the development data. Continuous variables were then scaled using means and SDs from the development data. Any missing variables were imputed as the mean of the development data (i.e., as 0 after scaling).

### S10. Example case

Consider a hypothetical 70 year male diagnosed with stage IV oral cavity SCC (see Supplementary Table S11 for full baseline characteristics). The conditional model predicts 80% probability of QoL decline in the next 2 years, given that the patient is alive. The joint model tells us that the probability of surviving and experiencing a QoL decline is 31%, and the predicted probability of death is 39%, which is almost 4-fold the mean observed mortality probability of 11% in the development cohort.

*Table S1 – List of candidate predictors and encoding used for developing the prediction models. All predictors are included in the extended set, while predictors included in the limited set are identified in the third column.*

| Predictor | Time | Included in the limited set | Parameters | Type |
|---|---|---|---|---|
| Sex | Diagnosis | yes | 1 | Binary |
| Age | Diagnosis | yes | 1 | Numeric |
| Education level | Diagnosis | yes | 1 | Ordinal |
| Marital status | Diagnosis | yes | 6 | Dummy |
| Regional deprivation index | Diagnosis | yes | 1 | Ordinal |
| Comorbidity index | Diagnosis | yes | 1 | Ordinal |
| Body mass index | Diagnosis | yes | 1 | Numeric |
| Primary tumour site (ICD code) | Diagnosis | yes | 6 | Dummy |
| Pre-treatment cTNM staging | Diagnosis | yes | 1 | Ordinal |
| HPV status | Diagnosis | yes | 3 | Dummy |
| Underwent Surgery | After treatment | yes | 1 | Binary |
| Received Chemotherapy | After treatment | yes | 1 | Binary |
| Received Radiotherapy | After treatment | yes | 1 | Binary |
| Smoking status | Diagnosis | yes | 4 | Dummy |
| Alcohol consumption | Diagnosis | yes | 1 | Numeric |
| EORTC QLQ C30 | Diagnosis, 4 months, 12 months | Only after treatment | 15 x 3 | Numeric |
| EORTC QLQ HN35 | Diagnosis, 4 months, 12 months | Only after treatment | 13 x 3 | Numeric |
| You and Cancer module[a] | 4 months, 12 months | No | 8 | Numeric |
| Your outlook module[a] | Diagnosis, 4 months, 12 months | No | 3 x 10 | Numeric |
| About you module[a] | Diagnosis | No | 6 | Numeric |

[a] – the detailed description of these predictors can be found in the Head & Neck 5000 study data manual available at https://headandneck5000.org.uk.

*Table S2 - Output from the pmsampsize R package for minimum sample size estimation for the limited set of parameters. Assumptions: 0.05 acceptable difference in apparent and adjusted R-squared; MMOE <= 1.1 in estimation of intercept and residual standard deviation. Minimum sample size required for new model development N = 1430.*

| Criteria | Samp_size | Shrinkage | Parameter | Rsq | SPP |
|---|---|---|---|---|---|
| **Criteria 1** | 1430 | 0.900 | 59 | 0.3 | 24.24 |
| **Criteria 2** | 827 | 0.840 | 59 | 0.3 | 14.02 |
| **Criteria 3** | 293 | 0.666 | 59 | 0.3 | 4.97 |
| **Criteria 4*** | 1430 | 0.900 | 59 | 0.3 | 24.24 |
| **Final** | 1430 | | | | |

SPP - Subjects per Predictor Parameter
* 95% CI for intercept = (72.9, 74.9), for sample size n = 1430

*Table S3 - Output from the pmsampsize R package for minimum sample size estimation for the extensive set of parameters. Assumptions: 0.05 acceptable difference in apparent and adjusted R-squared; MMOE <= 1.1 in* *estimation of intercept and residual standard deviation. Minimum sample size required for new model development N = 4249.*

| Criteria | Samp_size | Shrinkage | Parameter | Rsq | SPP |
|---|---|---|---|---|---|
| **Criteria 1** | 4249 | 0.900 | 171 | 0.3 | 24.85 |
| **Criteria 2** | 2395 | 0.836 | 171 | 0.3 | 14.01 |
| **Criteria 3** | 405 | 0.540 | 171 | 0.3 | 2.37 |
| **Criteria 4*** | 4249 | 0.900 | 171 | 0.3 | 24.85 |
| **Final** | 4249 | 0.900 | 171 | 0.3 | 24.85 |

SPP - Subjects per Predictor Parameter
* 95% CI for intercept = (73.3, 74.5), for sample size n = 4249

*Table S4 - TRIPOD Checklist: Prediction Model Development*

| ***Section/Topic*** | ***Item*** | ***Checklist Item*** | ***Page*** |
|---|---|---|---|
| ***Title and abstract*** | | | |
| *Title* | 1 | Identify the study as developing and/or validating a multivariable prediction model, the target population, and the outcome to be predicted. | Pg. 1 |
| *Abstract* | 2 | Provide a summary of objectives, study design, setting, participants, sample size, predictors, outcome, statistical analysis, results, and conclusions. | Pg. 1 |
| ***Introduction*** | | | |
| *Background and objectives* | 3a | Explain the medical context (including whether diagnostic or prognostic) and rationale for developing or validating the multivariable prediction model, including references to existing models. | Pg. 3 |
| | 3b | Specify the objectives, including whether the study describes the development or validation of the model or both. | Pg. 3 |
| ***Methods*** | | | |
| *Source of data* | 4a | Describe the study design or source of data (e.g., randomized trial, cohort, or registry data), separately for the development and validation data sets, if applicable. | Pg. 4, Supp.M. S7 and S8 |
| | 4b | Specify the key study dates, including start of accrual; end of accrual; and, if applicable, end of follow-up. | Supp. M. S7, Figure S1 |
| *Participants* | 5a | Specify key elements of the study setting (e.g., primary care, secondary care, general population) including number and location of centres. | Pg. 6 - 7, Supp. M. S7 |
| | 5b | Describe eligibility criteria for participants. | Supp. M. S1 |
| | 5c | Give details of treatments received, if relevant. | NA |
| *Outcome* | 6a | Clearly define the outcome that is predicted by the prediction model, including how and when assessed. | Pg. 5 |
| | 6b | Report any actions to blind assessment of the outcome to be predicted. | NA |
| *Predictors* | 7a | Clearly define all predictors used in developing or validating the multivariable prediction model, including how and when they were measured. | Pg. 5, Table S1 |
| | 7b | Report any actions to blind assessment of predictors for the outcome and other predictors. | NA |
| *Sample size* | 8 | Explain how the study size was arrived at. | Pg. 6, Table S2 & S3 |
| *Missing data* | 9 | Describe how missing data were handled (e.g., complete-case analysis, single imputation, multiple imputation) with details of any imputation method. | Pg. 6, Supp. M. S4 |
| *Statistical analysis methods* | 10a | Describe how predictors were handled in the analyses. | Pg. 6, Table S1 |
| | 10b | Specify type of model, all model-building procedures (including any predictor selection), and method for internal validation. | Pg. 6, Supp. M. S5 |
| | 10d | Specify all measures used to assess model performance and, if relevant, to compare multiple models. | Pg. 6 - 7, Supp. M. S6 |

| | | | |
|---|---|---|---|
| *Risk groups* | 11 | Provide details on how risk groups were created, if done. | NA |
| ***Results*** | | | |
| *Participants* | 13a | Describe the flow of participants through the study, including the number of participants with and without the outcome and, if applicable, a summary of the follow-up time. A diagram may be helpful. | Figure 1 and Supp. M. S1 |
| | 13b | Describe the characteristics of the participants (basic demographics, clinical features, available predictors), including the number of participants with missing data for predictors and outcome. | Supp. M. Table S5 |
| *Model development* | 14a | Specify the number of participants and outcome events in each analysis. | Pg. 7, Supp. M. Table S6 |
| | 14b | If done, report the unadjusted association between each candidate predictor and outcome. | NA |
| *Model specification* | 15a | Present the full prediction model to allow predictions for individuals (i.e., all regression coefficients, and model intercept or baseline survival at a given time point). | Supp. M. Tables S7, S8, and S9 |
| | 15b | Explain how to the use the prediction model. | Pg. 7 |
| *Model performance* | 16 | Report performance measures (with CIs) for the prediction model. | Pg. 8 |
| ***Discussion*** | | | |
| *Limitations* | 18 | Discuss any limitations of the study (such as nonrepresentative sample, few events per predictor, missing data). | Pg. 9 |
| *Interpretation* | 19b | Give an overall interpretation of the results, considering objectives, limitations, and results from similar studies, and other relevant evidence. | Pg. 9 - 10 |
| *Implications* | 20 | Discuss the potential clinical use of the model and implications for future research. | Pg. 10 |
| ***Other information*** | | | |
| *Supplementary information* | 21 | Provide information about the availability of supplementary resources, such as study protocol, Web calculator, and data sets. | Pg 6., Supp. M. Figure S2, |
| *Funding* | 22 | Give the source of funding and the role of the funders for the present study. | Pg. 11 |

*Table S5 – Table of characteristics for subjects alive at 36 Months from diagnosis with and without the QoL measurement.*

| **Characteristic** | **Total** N = 3172[1] | **Alive at 36 months and missing QoL** N = 1510[1] | **Alive at 36 months and not missing** N = 1662[1] |
|---|---|---|---|
| Sex | | | |
| Male | 2,361 (74) | 1,149 (76) | 1,212 (73) |
| Female | 811 (26) | 361 (24) | 450 (27) |
| Age | 61 (54, 67) | 60 (53, 67) | 61 (54, 68) |
| TNM Stage | | | |
| I | 793 (25) | 391 (26) | 402 (24) |
| II | 579 (18) | 292 (19) | 287 (17) |
| III | 437 (14) | 207 (14) | 230 (14) |
| IV | 1,354 (43) | 616 (41) | 738 (45) |
| Missing | 9 | 4 | 5 |
| Tumour location | | | |
| Oral cavity | 852 (27) | 415 (27) | 437 (26) |
| Oropharynx | 1,376 (43) | 587 (39) | 789 (47) |

| | | | |
|---|---|---|---|
| Nasopharynx | 62 (2.0) | 32 (2.1) | 30 (1.8) |
| Hypopharynx | 104 (3.3) | 58 (3.8) | 46 (2.8) |
| Larynx | 741 (23) | 401 (27) | 340 (20) |
| Nasal cavity/Sinuses | 37 (1.2) | 17 (1.1) | 20 (1.2) |
| Comorbidity index | | | |
| No co-morbidity | 1,439 (46) | 631 (43) | 808 (49) |
| Mild decompensation | 1,084 (35) | 502 (34) | 582 (36) |
| Moderate decompensation | 496 (16) | 287 (19) | 209 (13) |
| Severe decompensation | 93 (3.0) | 57 (3.9) | 36 (2.2) |
| Missing | 60 | 33 | 27 |
| HPV status | | | |
| Not obtained | 2,121 (67) | 1,061 (70) | 1,060 (64) |
| Negative | 237 (7.5) | 122 (8.1) | 115 (6.9) |
| Positive | 814 (26) | 327 (22) | 487 (29) |
| Household Income | | | |
| Less than £3999 | 87 (4.2) | 54 (6.8) | 33 (2.6) |
| £4000 - 7999 | 230 (11) | 117 (15) | 113 (8.8) |
| £8000 - 11999 | 272 (13) | 116 (15) | 156 (12) |
| £12000 - 17999 | 329 (16) | 136 (17) | 193 (15) |
| £18000 - 22999 | 207 (10.0) | 75 (9.4) | 132 (10) |
| £23000 - 28999 | 237 (11) | 87 (11) | 150 (12) |
| £29000 - 34999 | 168 (8.1) | 60 (7.5) | 108 (8.4) |
| £35000 or more | 549 (26) | 154 (19) | 395 (31) |
| Missing | 1,093 | 711 | 382 |
| Marital status | | | |
| Single | 286 (12) | 144 (15) | 142 (9.9) |
| Widowed | 148 (6.2) | 58 (6.1) | 90 (6.3) |
| Separated | 55 (2.3) | 34 (3.6) | 21 (1.5) |
| Divorced | 229 (9.6) | 105 (11) | 124 (8.6) |
| Married | 1,397 (58) | 489 (51) | 908 (63) |
| Living with a partner | 277 (12) | 126 (13) | 151 (11) |
| Missing | 780 | 554 | 226 |
| Education level | | | |

| | | | |
|---|---|---|---|
| Primary education | 27 (1.2) | 15 (1.7) | 12 (0.9) |
| Secondary education | 1,033 (45) | 463 (52) | 570 (41) |
| College | 803 (35) | 289 (32) | 514 (37) |
| High education | 415 (18) | 132 (15) | 283 (21) |
| Missing | 894 | 611 | 283 |
| Deprivation index (IMD10) | | | |
| Least deprived | 516 (19) | 184 (14) | 332 (23) |
| Less | 538 (20) | 224 (17) | 314 (22) |
| Middle | 613 (22) | 278 (22) | 335 (23) |
| More | 532 (19) | 261 (20) | 271 (19) |
| Most deprived | 551 (20) | 343 (27) | 208 (14) |
| Missing | 422 | 220 | 202 |
| BMI | 26 (23, 30) | 26 (23, 30) | 26 (24, 30) |
| Missing | 915 | 619 | 296 |
| Surgery | 1,516 (48) | 734 (49) | 782 (47) |
| Chemotherapy | 1,343 (42) | 593 (39) | 750 (45) |
| Radiotherapy | 2,408 (76) | 1,141 (76) | 1,267 (76) |
| Smoker status | | | |
| Never | 546 (24) | 163 (18) | 383 (28) |
| Former | 1,350 (59) | 536 (58) | 814 (59) |
| Current user | 404 (18) | 229 (25) | 175 (13) |
| Missing | 872 | 582 | 290 |
| Alcohol consumption | 21 (9, 36) | 22 (9, 45) | 21 (9, 36) |
| Missing | 1,362 | 800 | 562 |
| [1]n (%); Median (IQR) | | | |

*Table S6 – Table with the number of samples used for developing or for internal validating models, missing outcomes, and number of people that were deceased at the time of measurement.*

| Usage - Endpoint | N | Available | Events | Missing | Deceased |
|---|---|---|---|---|---|
| **Development - Survival** | 3572 | 3572 | 400 | 0 | 400 |
| **Development - Continuous QoL model** | 3572 | 1662 | N/A | 1510 | 400 |
| **Internal validation – QoL decline endpoint** | 3572 | 1351 | 238 | 1821 | 400 |
| **Interval validation – Joint model** | 3572 | 1751 | 638 | 1821 | 400 |

*Table S7 – Estimated parameters $\widehat{\beta_p} > 0$ for conformal LASSO with 5 models in the ensemble using the limited set of predictors. The description of each variable can be found in the data manual from H&N5000.*

| *Short description* | *Time* | *Model 1* | *Model 2* | *Model 3* | *Model 4* | *Model 5* |
|---|---|---|---|---|---|---|
| *Intercept* | - | 72.67143 | 73.58542 | 73.65172 | 73.08280 | 73.89246 |
| *TNM Stage* | Diagnosis | 0.17149 | 0.00000 | 0.40764 | 0.00000 | 0.07792 |
| *Comorbidity index* | Diagnosis | 0.00000 | -0.77995 | 0.00000 | 0.00000 | 0.00000 |
| *Household income* | Diagnosis | 1.58153 | 1.26044 | 1.13375 | 1.50845 | 1.18809 |
| *IMD10 wealth index* | Diagnosis | 0.00000 | -0.04946 | -0.09581 | 0.00000 | 0.00000 |
| *BMI* | Diagnosis | 0.00000 | 0.00000 | 0.00000 | 0.00000 | -0.11669 |
| *Alcohol consumption* | Diagnosis | 0.00000 | -0.39203 | 0.00000 | 0.00000 | 0.00000 |
| *C30 - Role functioning* | 12 Months | 0.07212 | 0.46001 | 0.43114 | 0.00000 | 0.00000 |
| *C30 - Physical functioning* | 12 Months | 3.44086 | 3.06238 | 3.25274 | 2.99327 | 4.03007 |
| *C30 - Emotional functioning* | 12 Months | 1.32868 | 0.19396 | 0.24664 | 0.00000 | 0.30052 |
| *C30 - Cognitive functioning* | 12 Months | 0.10860 | 0.72724 | 0.14644 | 0.18050 | 0.95732 |
| *C30 - Social functioning* | 12 Months | 0.00000 | 0.00000 | 0.06980 | 0.00000 | 0.00000 |
| *C30 - Fatigue* | 12 Months | -1.21368 | -1.62081 | -0.77422 | -0.74535 | -0.50734 |
| *C30 - Pain* | 12 Months | -1.86872 | -1.75943 | -1.96176 | -1.88418 | -2.43487 |
| *C30 - Dyspnoea* | 12 Months | -1.46195 | -0.67327 | -0.96852 | -1.03540 | -1.35993 |
| *C30 - Insomnia* | 12 Months | -0.19547 | -0.65975 | -0.59186 | -0.99989 | -1.14082 |
| *C30 - Constipation* | 12 Months | 0.00000 | -0.01273 | 0.00000 | 0.00000 | -0.23450 |
| *C30 - Diarrhoea* | 12 Months | 0.00000 | 0.00000 | 0.00000 | -0.27995 | -0.11196 |
| *C30 - GHS/QoL* | 12 Months | 6.00090 | 6.14884 | 6.36535 | 6.65971 | 6.37776 |
| *HN35 - Less sexuality* | 12 Months | -0.90220 | 0.00000 | -0.26708 | -1.02458 | 0.00000 |
| *HN35 - Feeling ill* | 12 Months | 0.00000 | 0.00000 | 0.00000 | -0.24317 | 0.00000 |
| *HN35 - Swallowing* | 12 Months | 0.00000 | 0.00000 | 0.00000 | -0.06008 | 0.00000 |
| *HN35 - Trouble with social eating* | 12 Months | 0.00000 | -0.18182 | -0.85838 | 0.00000 | 0.00000 |
| *HN35 - Teeth* | 12 Months | -0.09884 | -0.52902 | -0.45505 | -0.39362 | -0.04124 |
| *HN35 - Sticky saliva* | 12 Months | 0.00000 | -0.09782 | -0.08792 | 0.00000 | 0.00000 |
| *HN35 - Trouble with social contact* | 12 Months | 0.00000 | -0.36044 | -0.02354 | -0.29909 | 0.00000 |
| *HN35 - Cough* | 12 Months | 0.00000 | 0.00000 | 0.00000 | -0.00220 | 0.00000 |
| *Oropharynx* | Diagnosis | 0.59451 | 0.00000 | 0.00000 | 0.00000 | 0.00000 |
| *HPV status - Not obtained* | Diagnosis | 0.00000 | -0.12831 | -0.43961 | 0.00000 | -0.04024 |
| *HPV status - Positive* | Diagnosis | 0.00000 | 1.09683 | 0.00000 | 0.00000 | 0.00000 |
| *Marital status - Married* | Diagnosis | 0.97971 | 0.11113 | 0.82039 | 1.37860 | 0.00000 |

AY: About you module; YO: Your outlook module; YC: You and cancer module; C30: EORTC QLQ-C30 scales; HN35: EORTC QLQ-H&N35 scales

*Table S8 - Estimated parameters $\widehat{\beta_p} > 0$ for conformal LASSO with 5 models in the ensemble using the extensive set of predictors.*

| *Short description* | *Time* | *Model 1* | *Model 2* | *Model 3* | *Model 4* | *Model 5* |
|---|---|---|---|---|---|---|
| *Intercept* | - | 72.40275 | 73.28572 | 73.18158 | 72.77830 | 73.64123 |

| | | | | | | |
|---|---|---|---|---|---|---|
| *C30 - GHS/QoL* | 12 Months | 4.60903 | 4.25760 | 5.00654 | 4.85452 | 4.81959 |
| *C30 - Physical functioning* | Diagnosis | 3.21420 | 2.39072 | 1.94674 | 1.65303 | 2.89408 |
| *C30 - GHS/QoL* | 4 Months | 1.69303 | 1.66584 | 1.22930 | 2.41847 | 1.62391 |
| *Marital status - Married* | Diagnosis | 1.52369 | 0.58385 | 1.35771 | 1.68877 | 0.38729 |
| *C30 - Emotional func.* | 4 Months | 1.30504 | 1.18155 | 1.33287 | 1.74936 | 1.66434 |
| *Household income* | Diagnosis | 1.17071 | 0.86215 | 0.45689 | 1.11333 | 0.76871 |
| *C30 - Physical func.* | 12 Months | 1.15834 | 0.76065 | 1.57331 | 0.68042 | 1.30212 |
| *Oropharynx* | Diagnosis | 0.89847 | 0.00000 | 0.07635 | 0.26746 | 0.06848 |
| *C30 - GHS/QoL* | Diagnosis | 0.89598 | 1.34842 | 1.44801 | 1.13991 | 1.24268 |
| *YO - Friends* | 4 Months | 0.73619 | 0.33034 | 0.75377 | 1.03537 | 0.15900 |
| *C30 - Nausea* | 4 Months | 0.61952 | 0.30248 | 0.77766 | 1.19720 | 0.68667 |
| *YO - Keep busy* | 4 Months | 0.51039 | 0.39348 | 0.27397 | 0.74794 | 0.75386 |
| *C30 - Cognitive functioning* | 4 Months | 0.41985 | 0.45626 | 0.00005 | 0.00000 | 0.47144 |
| *C30 - Emotional functioning* | 12 Months | 0.40473 | 0.00000 | 0.00000 | 0.00000 | 0.00000 |
| *TNM Stage* | Diagnosis | 0.40289 | 0.20484 | 0.61477 | 0.21645 | 0.36378 |
| *C30 - QoL Summary* | Diagnosis | 0.38788 | 0.00000 | 0.00000 | 0.00000 | 0.00000 |
| *YO - Overall* | 4 Months | 0.31492 | 0.00000 | 0.00000 | 0.00000 | 0.00000 |
| *YC - How often* | 12 Months | 0.28591 | 0.25195 | 0.55972 | 0.57850 | 0.91924 |
| *YO - Overall* | Diagnosis | 0.18598 | 0.27730 | 0.10821 | 0.00000 | 0.49542 |
| *Appetite* | 4 Months | 0.16704 | 0.68347 | 0.69342 | 0.15788 | 0.31257 |
| *YO - Upset* | 4 Months | 0.11060 | 0.26719 | 0.00000 | 0.22744 | 0.00000 |
| *YO - Relax* | 4 Months | 0.00825 | 0.09724 | 0.03182 | 0.00000 | 0.46255 |
| *Comorbidity index* | Diagnosis | 0.00000 | -0.60195 | 0.00000 | 0.00000 | 0.00000 |
| *BMI* | 4 Months | 0.00000 | 0.00000 | 0.00000 | 0.00000 | -0.14683 |
| *Drink days* | 12 Months | 0.00000 | 0.00000 | 0.00000 | -0.20774 | 0.00000 |
| *Alcohol consumption* | Diagnosis | 0.00000 | -0.37861 | 0.00000 | 0.00000 | 0.00000 |
| *C30 - Cognitive functioning* | Diagnosis | 0.00000 | 0.00000 | 0.00000 | 0.76257 | 0.00000 |
| *C30 - Dyspnoea* | Diagnosis | 0.00000 | -0.08761 | -0.25837 | -0.04283 | -0.34795 |
| *C30 - Loss of appetite* | Diagnosis | 0.00000 | -0.26790 | 0.00000 | 0.00000 | 0.00000 |
| *C30 - Constipation* | Diagnosis | 0.00000 | 0.00000 | 0.01709 | 0.00000 | 0.00000 |
| *C30 - Role functioning* | 4 Months | 0.00000 | 0.00000 | 0.00000 | -0.11627 | 0.00000 |
| *C30 - Pain* | 4 Months | 0.00000 | -0.21039 | -0.76035 | -0.09656 | -0.64176 |
| *C30 - Dyspnoea* | 4 Months | 0.00000 | -0.36453 | -0.37448 | -0.52087 | 0.00000 |
| *C30 - Role functioning* | 12 Months | 0.00000 | 0.03796 | 0.00000 | 0.00000 | 0.00000 |
| *C30 - Cognitive functioning* | 12 Months | 0.00000 | 0.00000 | 0.00000 | 0.00000 | 0.05175 |
| *C30 - Social functioning* | 12 Months | 0.00000 | 0.00000 | 0.18149 | 0.00000 | 0.00000 |
| *C30 - Nausea or vomiting* | 12 Months | 0.00000 | 0.30716 | 0.00000 | 0.09964 | 0.00000 |
| *C30 - Insomnia* | 12 Months | 0.00000 | 0.00000 | 0.00000 | -0.14382 | -0.02092 |
| *C30 - Constipation* | 12 Months | 0.00000 | -0.13659 | -0.19646 | -0.01819 | -0.43882 |
| *C30 QoL Summary* | 12 Months | 0.00000 | 1.27715 | 0.00000 | 0.22738 | 1.68794 |
| *HN35 - Senses* | Diagnosis | 0.00000 | 0.00000 | -0.00986 | -0.02811 | 0.00000 |
| *HN35 - Openmouth* | Diagnosis | 0.00000 | 0.00000 | 0.00000 | 0.10315 | 0.07385 |
| *HN35 - Speech* | 4 Months | 0.00000 | 0.00000 | 0.35892 | 0.05728 | 0.14997 |

| | | | | | | |
|---|---|---|---|---|---|---|
| *HN35 - Swallow* | 4 Months | 0.00000 | 0.14717 | 0.00000 | 0.00000 | 0.00000 |
| *HN35 - Teeth* | 4 Months | 0.00000 | -0.51742 | -0.58802 | 0.00000 | -0.29163 |
| *HN35 - Saliva* | 4 Months | 0.00000 | 0.00000 | 0.00000 | 0.18214 | 0.06950 |
| *HN35 - Feeling ill* | 12 Months | 0.00000 | 0.00000 | 0.00000 | -0.10985 | 0.00000 |
| *HN35 - Teeth* | 12 Months | 0.00000 | -0.01240 | -0.04242 | -0.36539 | 0.00000 |
| *HN35 - Sense problems* | 12 Months | 0.00000 | 0.00000 | 0.00000 | 0.00767 | 0.00000 |
| *HN35 - Coughing* | 12 Months | 0.00000 | 0.00000 | -0.00689 | 0.00000 | 0.00000 |
| *YO - Upset* | Diagnosis | 0.00000 | 0.00000 | 0.07722 | 0.12610 | 0.00000 |
| *YO - Go wrong* | 4 Months | 0.00000 | 0.00000 | -0.09994 | -0.42191 | -0.28751 |
| *YO - Good things* | 4 Months | 0.00000 | 0.00000 | -0.19685 | 0.00000 | -0.33240 |
| *YO - Uncertain times* | 12 Months | 0.00000 | 0.00000 | 0.00000 | 0.00000 | -0.01183 |
| *AY - Mobility* | Diagnosis | 0.00000 | -0.37414 | -0.29874 | -0.49654 | -0.19009 |
| *AY - Self care* | Diagnosis | 0.00000 | -0.49285 | -0.39848 | -0.84854 | -0.08401 |
| *AY- Health State* | Diagnosis | 0.00000 | 0.00000 | 0.00000 | 0.14883 | 0.34713 |
| *YC - Recurrence* | 4 Months | 0.00000 | -0.22219 | -0.53505 | 0.00000 | 0.00000 |
| *YC - Strong feel* | 4 Months | 0.00000 | -0.02227 | 0.00000 | 0.00000 | 0.00000 |
| *YC - Strongical feeltioning* | 12 Months | 0.00000 | 0.05701 | 0.00000 | 0.18207 | 0.00000 |
| *HPV status - Not obtained* | Diagnosis | 0.00000 | 0.00000 | -0.15841 | 0.00000 | 0.00000 |
| *HPV status - Positive* | Diagnosis | 0.00000 | 0.90206 | 0.00000 | 0.00000 | 0.00000 |
| *HN35 - Cough* | 4 Months | -0.02677 | 0.00000 | 0.00000 | -0.38787 | -0.00910 |
| *AY - Health usual activities* | Diagnosis | -0.03700 | 0.00000 | 0.00000 | 0.00000 | 0.00000 |
| *YO - Good Things* | Diagnosis | -0.04325 | 0.00000 | 0.00000 | 0.00000 | 0.00000 |
| *HN35 - Trouble with social eating* | 12 Months | -0.06142 | -0.03624 | -0.92583 | 0.00000 | 0.00000 |
| *HN35 - Trouble with social contact* | 12 Months | -0.10711 | -0.60485 | -0.36622 | -0.76334 | 0.00000 |
| *YO - Go my way* | 4 Months | -0.11362 | -0.40753 | 0.00000 | 0.00000 | 0.00000 |
| *HN35 - Cough* | Diagnosis | -0.21257 | -0.01164 | -0.08675 | -0.61717 | 0.00000 |
| *C30 - Insomnia* | 4 Months | -0.22814 | -0.06318 | 0.00000 | 0.00000 | 0.00000 |
| *HN35 - Teeth problems* | Diagnosis | -0.30950 | -0.10053 | -0.54778 | -0.51056 | -0.29365 |
| *HN35 - Dry mouth* | Diagnosis | -0.33883 | -0.35078 | -0.02403 | 0.00000 | 0.00000 |
| *YO - Go wrong feeling* | Diagnosis | -0.50543 | -0.16644 | -0.63271 | -0.04671 | -0.06118 |
| *C30 - Fatigue* | Diagnosis | -0.52470 | -0.94817 | -1.05375 | -0.01576 | -0.30203 |
| *HN35 - Less sexuality* | 12 Months | -0.71934 | 0.00000 | 0.00000 | -0.73221 | 0.00000 |
| *C30 - Dyspnoea* | 12 Months | -0.97630 | 0.00000 | 0.00000 | -0.34208 | -0.55482 |
| *C30 -Fatigue* | 12 Months | -1.17170 | -1.55773 | -1.22854 | -0.95940 | -0.50462 |
| *C30 - Pain* | 12 Months | -1.39649 | -1.40490 | -1.24959 | -1.59834 | -1.68185 |

AY: About you module; YO: Your outlook module; YC: You and cancer module; C30: EORTC QLQ-C30 scales; HN35: EORTC QLQ-H&N35 scales.

*Table S9 – Logistic regression with L1 penalty model coefficients, mean, and variance for variables standardization.*

| *Short description* | Parameter | Mean | Variance |
|---|---|---|---|
| *Intercept* | -2.307169115 | - | - |
| *Age* | 0.4542332257903460 | 61.0831467 | 113.8293128 |
| *TNM stage* | 0.26693079863428600 | 2.772944148 | 1.525487459 |

| | | | |
|---|---|---|---|
| *Comorbidity index* | 0.32038177542516200 | 1.809591778 | 0.734798092 |
| *Household income* | 0. 00000000000000000 | 5.0683391 | 5.15103911 |
| *Education level* | 0. 00000000000000000 | 2.692064745 | 0.594477103 |
| *Deprivation Index (IMD10)* | 0.0876692270000000 | 2.055680721 | 1.950230845 |
| *BMI* | -0.1400950520000000 | 26.76861919 | 28.55096723 |
| *Alcohol consumption* | 0.09094695700000000 | 29.56802383 | 654.3384711 |
| *C30 - role functioning* | 0. 00000000000000000 | 77.55830486 | 806.725255 |
| *C30 - physical functioning* | -0.02058125100000000 | 79.10557407 | 515.9651832 |
| *C30 - emotional functioning* | 0.10850434764139000 | 76.25711469 | 596.7356058 |
| *C30 - cognitive functioning* | 0.11257168676948300 | 79.3942256 | 518.8525912 |
| *C30 - social functioning* | -0.02071268500000000 | 75.2088253 | 795.169204 |
| *C30 - fatigue* | 0. 00000000000000000 | 33.44349846 | 683.8965152 |
| *C30 - nausea* | 0.024193270711295200 | 6.610143114 | 230.5510472 |
| *C30 - pain* | 0. 00000000000000000 | 21.97220894 | 693.9944586 |
| *C30 - dyspnoea* | 0.03082531000000000 | 17.85097904 | 744.8718322 |
| *C30 - insomnia* | -0.09476975100000000 | 31.18935617 | 1007.922858 |
| *C30 - appetite* | 0.029162573494252900 | 24.21234881 | 1053.648592 |
| *C30 - constipation* | 0.011321072992426000 | 15.74334776 | 670.2480879 |
| *C30 - diarrhoea* | 0.03918856000000000 | 6.638483875 | 308.730443 |
| *C30 – GHS/QoL* | 0. 00000000000000000 | 68.99279449 | 500.9331517 |
| *HN35 - pain* | 0. 00000000000000000 | 18.25281777 | 420.1223371 |
| *HN35 - speech* | 0. 00000000000000000 | 20.11997614 | 514.8894751 |
| *HN35 - less sexuality* | 0.04301072600000000 | 39.03695756 | 1439.572425 |
| *HN35 - dry mouth* | 0. 00000000000000000 | 51.16023732 | 1303.92195 |
| *HN35 - feeling ill* | 0.10761370566843600 | 14.39358142 | 538.9838641 |
| *HN35 - swallowing* | 0.09563695600000000 | 16.65868431 | 471.4106373 |
| *HN35 - trouble with social eating* | 0.22733167097258300 | 24.87986519 | 820.3710612 |
| *HN35 - teeth* | 0.04978162900000000 | 20.99712319 | 961.0596239 |
| *HN35 - sticky saliva* | -0.02047082100000000 | 37.9060225 | 1319.361689 |
| *HN35 - trouble with social contact* | -0.02622036700000000 | 25.75900535 | 740.5688344 |
| *HN35 - open mouth* | -0.02878522200000000 | 11.68424404 | 401.2860403 |
| *HN35 - cough* | 0.026336622655421700 | 19.01176422 | 843.2586597 |
| *Sex - male* | 0. 00000000000000000 | 30.16171986 | 726.8435484 |
| *ICD group - oral cavity* | -0.37247650000000000 | - | - |
| *ICD group - oropharynx* | 0.052630503000000000 | - | - |
| *ICD group - nasopharynx* | -0.28932942400000000 | - | - |
| *ICD group - hypopharynx* | 0. 00000000000000000 | - | - |
| *ICD group - larynx* | 0. 00000000000000000 | - | - |
| *ICD group - nasal cavity/sinuses* | 0. 00000000000000000 | - | - |
| *HPV status - negative* | 0. 00000000000000000 | - | - |
| *HPV status - not obtained* | 0. 00000000000000000 | - | - |
| *HPV status - positive* | 0. 00000000000000000 | - | - |
| *Marital status - single* | -0.28083891700000000 | - | - |
| *Marital status - widowed* | 0. 00000000000000000 | - | - |
| *Marital status - separated* | 0. 00000000000000000 | - | - |
| *Marital status - married* | 0. 00000000000000000 | - | - |
| *Marital status - divorced* | -0.18123684200000000 | - | - |
| *Marital status - living with a partner* | 0. 00000000000000000 | - | - |
| *Marital status - missing* | -0.01082154300000000 | - | - |
| *Surgery* | 0.09267789500000000 | - | - |
| *Chemotherapy* | 0. 00000000000000000 | - | - |
| *Radiotherapy* | 0.04981126105838280 | - | - |
| *Tobacco use - current user* | 0.34615288604548100 | - | - |
| *Tobacco use - former* | 0.39188278698427500 | - | - |
| *Tobacco use - never* | -0.1113631430000000 | - | - |
| *Tobacco use - missing* | -0.3403354300000000 | - | - |

*Table S10 – Extra performance assessment for LASSO model developed with limited set of predictors. Model performance by groups of patients classified with high propensity score (similar characteristics to patients with missing outcome) and low propensity score.*

| | *C-index [CI 95%]* |
|---|---|
| *Overall* | 0.656 [0.61, 0.69] |
| *High propensity score* | 0.662 [0.55, 0.73] |
| *Low propensity score* | 0.651 [0.60, 0.69] |

*Table S11 – Table of characteristics for the data used in external validation. The data was compiled from 3 studies conducted in Germany (UMM1, 2 and 3) and one study from Italy (INT).*

| **Characteristic** | **Overall**<br>N = 497[1] |
|---|---|
| Study | |
| INT | 56 (11) |
| UMM1 | 105 (21) |
| UMM2 | 230 (46) |
| UMM3 | 106 (21) |
| Age | 62 (54, 69) |
| Missing | 32 |
| Sex | |
| Male | 434 (87) |
| Female | 63 (13) |
| Marital status | |
| Single | 67 (17) |
| Married/Living with a partner | 268 (66) |
| Divorced/Separated | 48 (12) |
| Widowed | 22 (5.4) |
| Missing | 92 |
| BMI | 0 (NA) |
| Missing | 497 |
| Deprivation index (IMD10) | 0 (NA) |
| Missing | 497 |
| TNM Stage 7th Ed. | |
| I | 147 (32) |
| II | 78 (17) |
| III | 71 (15) |

| | |
|---|---|
| IV | 165 (36) |
| Missing | 36 |
| Tumour location | |
| Oral cavity | 63 (13) |
| Oropharynx | 36 (7.2) |
| Hypopharynx | 40 (8.0) |
| Larynx | 334 (67) |
| Nasal cavity and paranasal sinuses | 3 (0.6) |
| Overlapping several areas | 21 (4.2) |
| Radiotherapy | |
| Yes | 243 (56) |
| No | 192 (44) |
| Missing | 62 |
| Chemotherapy | |
| Yes | 111 (26) |
| No | 317 (74) |
| Missing | 69 |
| Surgery | |
| Yes | 257 (53) |
| No | 232 (47) |
| Missing | 8 |
| Smoker status | |
| Current | 91 (31) |
| Former | 157 (54) |
| Never | 45 (15) |
| Missing | 204 |
| Alcohol consumption | 17 (1, 42) |
| Missing | 246 |
| Education level | |
| Low | 165 (42) |
| Medium | 139 (35) |
| High | 93 (23) |

| | |
|---|---|
| Missing | 100 |
| Household income (EUR/month) | |
| Less than 500 | 19 (7.6) |
| 500 - 1500 | 92 (37) |
| 1501 - 2500 | 95 (38) |
| 2501 - 3500 | 33 (13) |
| More than 3500 | 12 (4.8) |
| Missing | 246 |
| QLQ-C30 GHS/QoL - Baseline | 58 (41, 75) |
| Missing | 99 |
| QLQ-C30 GHS/QoL - Outcome | 66 (50, 83) |
| Comorbidity index | 0 (NA) |
| Missing | 497 |
| HPV Status | |
| Not obtained | 497 (100) |
| QoL Decline | 79 (20) |
| Missing | 99 |

[1]n (%); Median (IQR)

*Table S12 - Table of characteristics for the data used in external validation of survival submodel. The data is from the BD4Decide study.*

| | Overall<br>(N=281) |
|---|---|
| **Clinical age at diagnosis** | |
| Mean (SD) | 61.4 (11.2) |
| Median [Min, Max] | 61.0 [21.0, 93.0] |
| **Clinical sex** | |
| Female | 81 (28.8%) |
| Male | 200 (71.2%) |
| **ctn stage at diagnosis** | |
| Stage III | 60 (21.4%) |
| Stage IVA | 197 (70.1%) |
| Stage IVB | 24 (8.5%) |
| **Tumor region** | |
| Hypopharynx | 32 (11.4%) |
| Larynx | 41 (14.6%) |
| Oral Cavity | 83 (29.5%) |
| Oropharynx | 125 (44.5%) |
| **Overall comorbidity score** | |
| 0 | 82 (29.2%) |
| 1 | 123 (43.8%) |
| 2 | 65 (23.1%) |
| 3 | 11 (3.9%) |
| **HPV status** | |
| Negative | 39 (13.9%) |
| Positive | 86 (30.6%) |
| Missing | 156 (55.5%) |
| **Number of alcohol units per day** | |
| Mean (SD) | 2.77 (4.68) |
| Median [Min, Max] | 0 [0, 32.2] |
| Missing | 4 (1.4%) |
| **Alcohol at time of diagnosis** | |
| Current | 124 (44.1%) |
| Former | 20 (7.1%) |
| Never | 133 (47.3%) |
| Unknown | 4 (1.4%) |
| **Smoker at time of diagnosis** | |
| Current | 134 (47.7%) |
| Former | 70 (24.9%) |
| Never | 75 (26.7%) |
| Unknown | 2 (0.7%) |
| **Undergone cancer surgery** | |
| No | 107 (38.1%) |
| Yes | 174 (61.9%) |
| **Chemotherapy treatment** | |
| No | 93 (33.1%) |
| Yes | 188 (66.9%) |
| **Radiotherapy treatment** | |
| No | 28 (10.0%) |
| Yes | 253 (90.0%) |

*Table S13 – Table with the characteristics of the example case*

| Characteristics | Value |
|---|---|
| **Marital Status** | Single |
| **Education Level** | Maximum 6 years |
| **Tobacco Use** | Current user |
| **Household Income** | Less than 3999 GBP |
| **Age at Diagnosis** | 70 |
| **Body Mass Index (BMI)** | 30 |

| | |
|---|---|
| **Alcohol consumption** | 77 |
| **Tumour region** | Oral cavity |
| **Deprivation index (IMD10)** | Most deprived |
| **Sex** | Male |
| **Comorbidity Index** | Severe |
| **HPV Status** | Not obtained |
| **TNM Stage** | IV |
| **Chemotherapy** | No |
| **Radiotherapy** | Yes |
| **Surgery** | Yes |
| **EORTC QLQ-C30 Appetite Loss** | 39 |
| **EORTC QLQ-C30 Cognitive Function** | 60 |
| **EORTC QLQ-C30 Constipation** | 33 |
| **EORTC QLQ-C30 Diarrhoea** | 13 |
| **EORTC QLQ-C30 Dyspnoea** | 46 |
| **EORTC QLQ-C30 Emotional Function** | 64 |
| **EORTC QLQ-C30 Fatigue** | 55 |
| **EORTC QLQ-C30 Global Health Status** | 88 |
| **EORTC QLQ-C30 Insomnia** | 46 |
| **EORTC QLQ-C30 Nausea/Vomiting** | 6 |
| **EORTC QLQ-C30 Pain** | 49 |
| **EORTC QLQ-C30 Physical Function** | 52 |
| **EORTC QLQ-C30 Role Function** | 56 |
| **EORTC QLQ-C30 Social Function** | 59 |
| **EORTC QLQ-HN35 Coughing** | 46 |
| **EORTC QLQ-HN35 Dry Mouth** | 66 |
| **EORTC QLQ-HN35 Ill Feeling** | 39 |
| **EORTC QLQ-HN35 Trouble Opening Mouth** | 33 |
| **EORTC QLQ-HN35 Pain** | 29 |
| **EORTC QLQ-HN35 Saliva** | 66 |
| **EORTC QLQ-HN35 Senses Problems** | 36 |
| **EORTC QLQ-HN35 Sexual Problems** | 59 |

| | |
|---|---|
| **EORTC QLQ-HN35 Social Contact Problems** | 14 |
| **EORTC QLQ-HN35 Social Eating Problems** | 45 |
| **EORTC QLQ-HN35 Speech Problems** | 37 |
| **EORTC QLQ-HN35 Swallowing Problems** | 36 |
| **EORTC QLQ-HN35 Teeth Problems** | 19 |

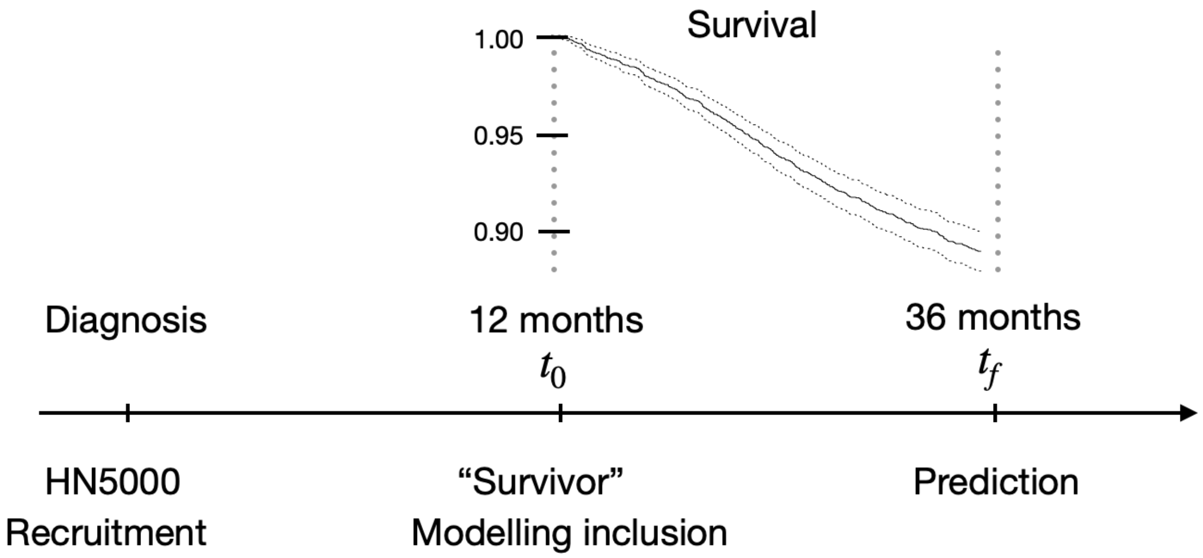


*Figure S1 – The figure shows the timeline for patients included in the model development superposed with the Kaplan-Meier curve. Patients were recruited to the HN5000 study at diagnosis/before treatment. Patients that were alive and disease free at 12 months after diagnosis were eligible for model development. The prediction is done at 36 months after diagnosis.*

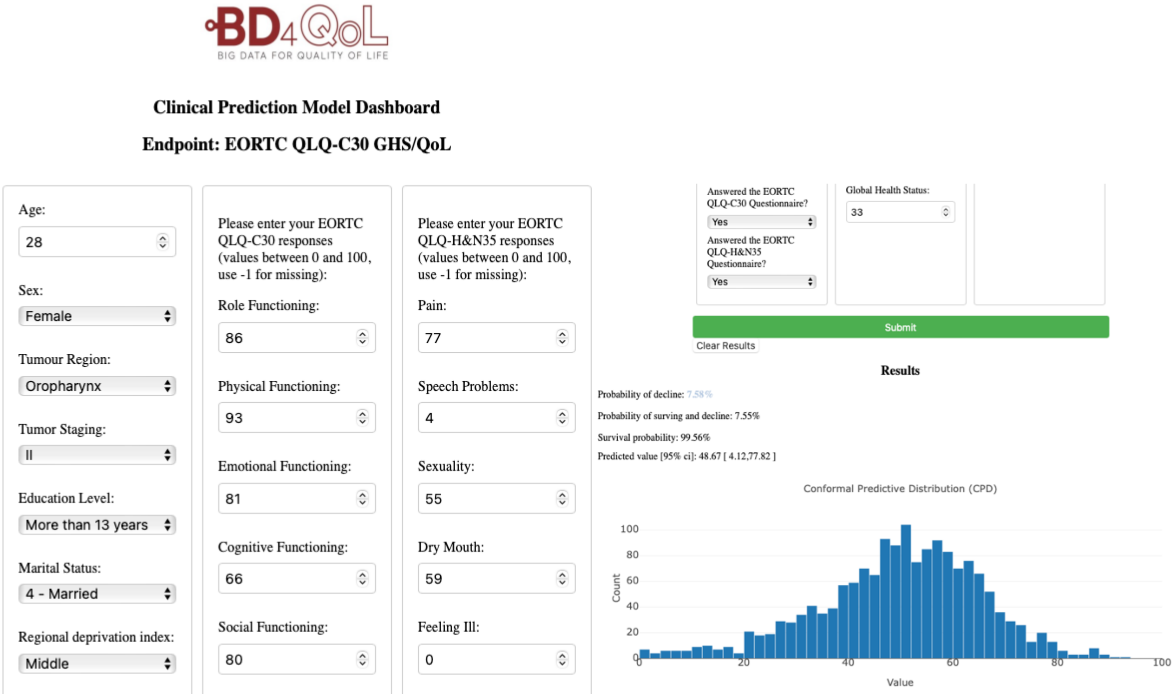


*Figure S2 –The model can be used via the BD4Predict dashboard (https://ocbe-uio.github.io/bd4predict) which requires no technical knowledge. Users can provide patient data via (left) dashboard and retrieve (right) predictions from the model easily. Advanced users can interact with the models programmatically via API (https://bd4predict.azurewebsites.net/docs), documented in SWAGGER and REDOC standards.*

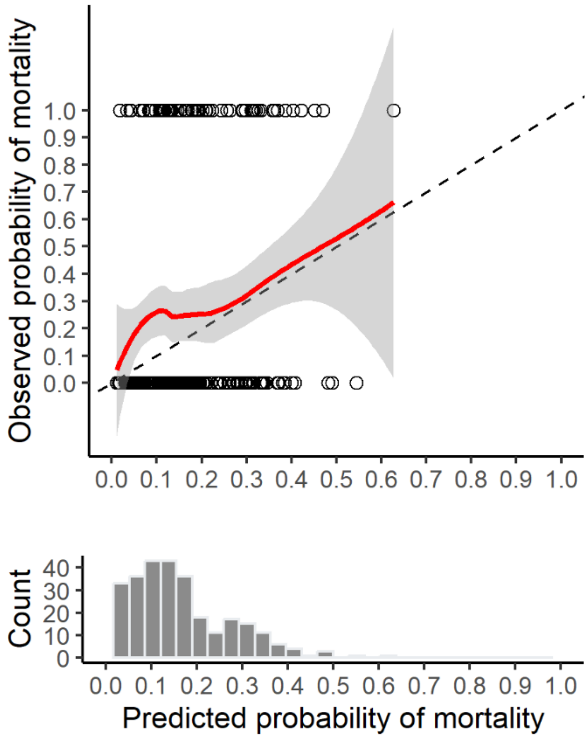


*Figure S3 - Calibration curve showing the relationship between predicted probability of mortality from the survival submodel and the observed mortality in BD2Decide data. Dotted line shows the 1:1 line, the red line is a smoothed line with a 95% confidence interval in grey.*

1. Compton SEC, Cancer AJC on, others. the 7th edition of the AJCC cancer staging manual and the future of TNM Ann. Surg Oncol. 17:1471–4.
